# A model-based evaluation of the cost-effectiveness of paediatric and elderly vaccination against pneumococcal infection in England

**DOI:** 10.64898/2026.02.26.26347158

**Authors:** Matt J. Keeling, Omar El Deeb, Phuong Bich Tran, Stavros Petrou, Edward M. Hill

## Abstract

Infection with pnuemococcus bacteria is generally mild but can be more severe in the young and elderly, causing invasive pneumococcal disease (IPD) and community-acquired pneumonia (CAP). Although paediatric pneumococcal conjugate vaccine (PCV) programmes and elderly pneumococcal polysaccharide vaccine (PPV) programmes have reduced cases, we estimate that pneumococcal infection still leads to direct health care costs of around £68M and approximately 16 thousand QALY losses in England per year. The public health situation is complicated by the large number of interacting serotypes, such that while serotype-specific vaccines reduced the target serotypes others arose to replace them.

Here we develop a novel (relatively) low-dimensional model to capture the interaction of 26 common pneumococcal serotypes. The model is matched to English IPD data from 2000-2023 and to five carriage studies (conducted in 2001/02, 2008, 2012, 2015 and 2018). When combined with a health economic approach, this model allows us to calculate the willingness to pay for paediatric vaccination with PCV7 (introduced in England in 2006), PCV13 (introduced in England in 2010) and the future vaccination of both the young and elderly with PCV20, which offers protection against 20 serotypes.

Due to rapid serotype replacement, we find that the introduction of PCV7 vaccination in 2006 was not cost effective - a result that could not have been anticipated at the time, but is supported by simple statistical fits to the IPD data. In contrast, switching to PCV13 in 2010 and switching to PCV20 in 2026 are both associated with a high willingness to pay for a single dose. Given pneumococcal disease has shifted over time to become predominantly in the older adult population, we find that switching from PPV23 to PCV20 vaccination in those aged 65 and introducing an additional PCV20 vaccine at age 75 are both cost effective for a sufficiently low vaccine price.

The inference that underpins our model is unfortunately limited by the available data and the high-dimensional nature of multiple interacting serotypes. Future sampling of carriage from older adults would greatly improve our confidence, as would national estimates of CAP.

## 1 Introduction

*Streptococcus pneumoniae* (or pneumococcus) bacteria can cause many types of infection: from mild carriage to severe invasive pneumococcal disease (IPD) with multiple sequelae. Spread of this bacteria is generally through direct contact with respiratory secretions [1]. The symptoms and more severe complications experienced from an infection depend on the part of the body that is infected. IPD is the most severe form, when the bacteria has spread to the lungs (causing pneumonia), bloodstream (causing bacteraemia) and/or the lining of the brain and spinal cord (causing meningitis). IPD is a notable cause of morbidity and mortality worldwide (an estimated 1.6 million deaths globally [2]), with the highest incidence typically observed among young children and older individuals [3]. Mucosal infections can lead to Otitis media (middle ear infection) and sinusitis. Community-acquired pneumonia (CAP) is a respiratory disease that can be caused by pneumococcus bacteria, although other causes are also common; we use the term pneumococcal community-acquired pneumonia (pCAP) to distinguish cases caused by pneumococcus bacteria. While pCAP is more common than IPD, the outcomes are usually less severe.

The key public health intervention to tackle pneumococcal infection and reduce disease burden has been administering pneumococcal conjugate vaccines (PCVs) to children, as recommended by WHO [4], and pneumococcal polysaccharide vaccines (PPV) to older adults. PCVs have been highly effective in preventing invasive pneumococcal disease in children. PCVs can also provide protection against carriage [5], meaning the protection provided to children can impact on the risk of infection across all age-groups.

The UK introduced the seven-valent PCV7 for infants in 2006, with primary doses at two months and four months of age, followed by a booster at 12–13 months (we refer to this as a 2+1 schedule). In April 2010, PCV7 was replaced with 13-valent PCV (PCV13), which covered an additional six serotypes. In January 2020, the UK moved from a 2+1 to a 1+1 schedule (primary dose at 12 weeks of age; booster at 1 year of age), at a substantial reduction in vaccine costs but little, if any, expected decline in protection [6] (though any epidemiological signatures of this change is likely to be masked by the impact of the COVID-19 pandemic). These vaccination programmes have seen a reduction in vaccine-type IPD cases, especially in children (Fig. 1). In England, cases of IPD in children under 2 have reduced from around 600 per year pre-2006 (before the introduction of the pneumococcal vaccine programme for infants) to just 156 in 2022. However, an unintended consequence has been serotype replacement, with non-vaccine serotypes (NVTs) having increased among asymptomatic carriers and IPD cases. The increasing prevalence of NVTs has particularly impacted those aged 65 years and over; increases in non-vaccine IPD in this age-group after the introduction of PCV13, which has largely negated the reduction in vaccine-type IPD.

**Fig. 1.**
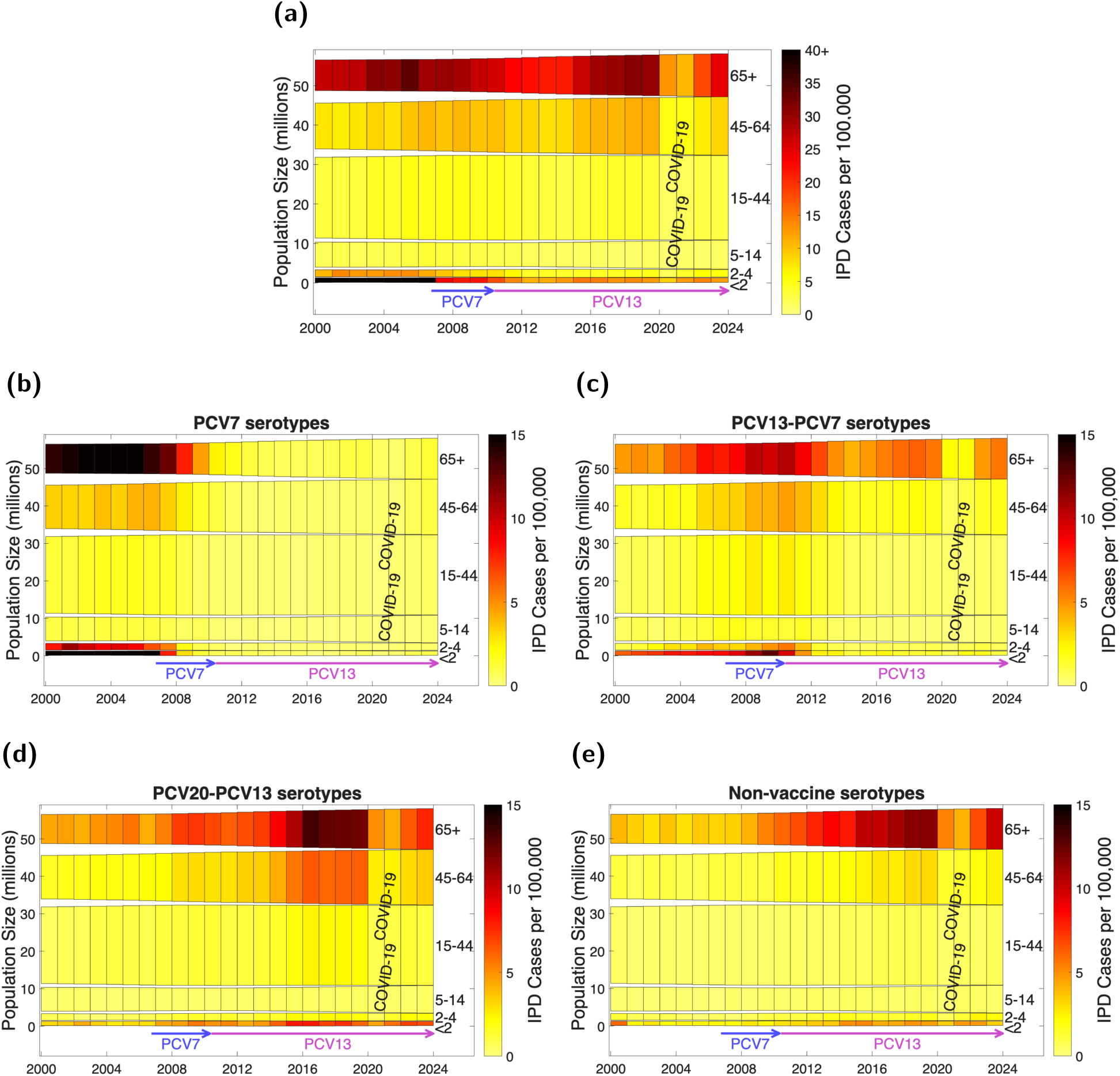
Change in the number of IPD cases recorded per 100,000 in six age-groups. Bar heights correspond to the population size of the associated age group, which change over time. Bar shading depicts the number of IPD cases per 100,000 for the specified age group per calendar year for: **(a)** all serotypes; **(b)** serotypes contained in the PCV7 vaccine (4, 6B, 9V, 14, 18C, 19F, 23F); **(c)** the six additional serotypes contained in the PCV13 vaccine; (1, 3, 5, 6A, 7F, 19A) **(d)** the seven additional serotypes contained in the PCV20 vaccine (8, 10A, 11A, 12F, 15B, 22F, 33F); **(e)** Serotypes not contained in PCV7, PCV13 or PCV20 vaccines. Arrows denote when PCV7 and PCV13 vaccines became part of the infant vaccination programme. We also observe a decrease in reported cases during the 2020-2021 COVID-19 due to the impact of pandemic controls on transmission.

Non-pharmaceutical interventions (NPIs) focused on control of the COVID-19 pandemic also disrupted the transmission of many other pathogens. Pneumococcal infections were included in this disruption to pathogen circulation, leading to a reduction in recorded IPD cases in 2020 and 2021 [7]. Post-COVID, surveillance data from 2022-2023 reports most cases of IPD in England being caused by non-PCV13 serotypes or serotype 3 [8] (PCV vaccines are known to provide low protection against serotype 3 [9]). The development of higher valency PCVs that cover some of the most common replacing serotypes (e.g. serotypes 8, 9N, 10A, 11A, 22F or 33F) could therefore potentially limit the future incidence of IPD - although serotype replacement could again weaken the impact. One such vaccine is the 20-valent PCV (PCV20, which covers an additional seven serotypes); this has been licensed for paediatric use by the European Medicines Agency [10].

Changes in vaccination programmes can come at a considerable cost, as well as epidemiological implications. Combined epidemiological and health economic models are therefore an important tool to assess whether any change in a vaccination programme is cost-effective. That is, whether the value placed on predicted health benefits or improvements in social welfare combined with reduction in treatment costs, outweigh the incremental cost associated with the change in programme. Such analyses have been performed in the context of the paediatric pneumococcal vaccination programme for England, both for the introduction of PCV7 [11] and investigating the cost effectiveness of a transition from PCV7 to PCV13 [12]. These earlier results suggest that the introduction of PCV7 would be cost-effective at £10-15 (for 3 doses), and that in 2010 PCV13 would be cost-effective at £49.60 (plus £7.50 administration fees) compared to stopping infant vaccination (again for 3 doses).

Given the dynamic nature of pneumococcal serotypes and ongoing serotype replacement (Fig. 1), forecasting the long-term cost-effectiveness of switching to PCV20 immunisation programme is crucial for informing public health decision-making. The potential epidemiological implications of a transition to PCV20 from PCV13, has been investigated in a modelling study by Choi *et al.* [13]. The authors projected the switch to PCV20 would reduce overall IPD due to the relatively high severity of the additional seven serotypes covered by PCV20 (8, 10A, 11A, 12F, 15B, 22F and 33F) compared to non-vaccine serotypes that could replace them. A cost-effectiveness study by Pfizer used a dynamic transmission model, finding for a 5-year time horizon that PCV20 (1+1 schedule) was cost-saving compared with PCV13 (1+1 schedule) [14]. Outside of the UK, another study by Pfizer performed a cost-effectiveness of a PCV20 catch-up program in US children aged 14–59 months who completed the previously recommended PCV13 (3+1) series [15]; assessed over a 10-year time horizon, they found such an infant PCV20 catch-up campaign was estimated to further reduce pneumococcal disease and associated healthcare system and societal costs, and to be cost neutral.

In parallel to the updates to the paediatric pneumococcal vaccination programme with higher-valency PCVs, there is an increasing policy focus on the vaccine used in the adult pneumococcal vaccination programme. Vaccination of older adults began in 2003, and from 2005 onwards all individuals aged 65 or over have been eligible for a single dose of PPV23. The UK’s Joint Committee on Vaccination and Immunisation (JCVI) agreed during 2023 that either PCV20 or PPV23 could be used for the adult pneumococcal vaccination programme [16]. A static cohort modelling study in 2022 [17] found that replacing PPV23 with PCV20 for adults above 65 years (and adults aged 18-64 years with underlying conditions) in England at vaccine list prices was cost saving in 85% of model simulations. A more recent cost-effectiveness study of pneumococcal immunisation strategies for the elderly in England [18] showed that the use of either PPV23 or PCV20 for the adult pneumococcal vaccination programme were likely to be cost-effective. These cost-effectiveness outcomes were again based on a static cohort model with probabilistic sensitivity analysis and assumed the cost of vaccine administration was £10.06, the list price for PPV23 was £16.80 and the list price for PCV20 was £56.80. The possibility of a transition from PPV23 to PCV20 in adults aged 65 and older in England is therefore of additional interest.

Here we aim to capture the dynamics of pneumococcal infection over time, its response to the introduction of PCV7 and PCV13, and the number of IPD cases recorded in each age-group. By capturing the observed dynamics, we are able to project future dynamics where other changes are introduced. The hospital data collated by UKHSA contains 105 different serotypes (although 80% of IPD cases are associated with the 33 most common serotypes). Given multiple infections are relatively common, a standard SIRS-type modelling framework would need to account for the number of individuals susceptible to, infected with, or recovered from each serotype - leading to over five quadrillion (5.559 × 10^15^) equations. We instead adopt a formulation that considers the number of hosts susceptible to each serotype [19, 20] and captures the interaction between serotypes through independence assumptions. As such, we are able to capture 26 serotypes, nine age groups and three vaccine types with just 3042 equations.

The applied public health novelty of this modelling comes from our ability to track a wide spectrum of serotypes and their cross-reaction; rather than the common approach to date which has treated multiple serotypes as a single unit (e.g. all serotypes in PCV7, or all serotypes in PCV13 but not in the PCV7 vaccine, [12, 13]). Applying our novel approach to England, we fit the model to pneumococcal carriage [21, 22] and IPD outcome data (provided by the UK Health Security Agency) to capture the herd immunity and serotype replacement precipitated by the PCV7 and PCV13 paediatric pneumococcal vaccination programmes in England. We then evaluated the willingness to pay for changing from PCV13 to PCV20 in infants but maintaining the current UK 1+1 pneumococcal vaccine dose schedule. This evaluation is made more complex by the potential switch from PPV23 to PCV20 in those aged 65. As part of our scenario assessment, we therefore considered different combinations of pneumococcal vaccine products being used from 2026 onward for infants (PCV13 or PCV20) and adults (PPV23 or PCV20), respectively. We find the cost-effectiveness of replacing PCV13 by PCV20 in infants to be especially sensitive to the time-horizon and PCV20 effectiveness. Over long time-horizons (50 years) the switch to PCV20 could be cost-effective provided there was not a substantial decline in its efficacy against carriage and disease. We find the switch from PPV23 to PCV20 in older adults is far less cost-effective but still has a positive willingness to pay. Finally, given the dominance of IPD in the elderly, we consider an additional dose of PCV20 at age 75, and again show there is a relatively high willingness to pay per dose. In most cases, the willingness to pay is largely unaffected if standard uncertainty assessment protocols are applied [23].

## 2 Methods

Our model-based evaluation of the cost-effectiveness of paediatric and elderly vaccination against pneumococcal infection in England required a combination of data analysis, infectious disease modelling and health economics. We begin by describing the multiple data sources that are used throughout this paper (Section 2.1), the structure of our pneumococcal transmission model and the inference procedure used to fit the model to the available epidemiological data (Section 2.2), the health economic model (Section 2.3), how we calculated willingness to pay (WTP) thresholds in our cost-effectiveness analysis (Section 2.4) and the vaccination programme scenarios that we consider (Section 2.5).

### 2.1 Data sources

Any applied epidemiological model relies on high-quality data. For the long-term behaviour of pneumococcal infection and disease, we require an estimate of population demographics, vaccine uptake, vaccine effectiveness, carriage of infection and severe disease (IPD, pCAP and associated sequelae), which we briefly describe below. We consider in some detail the dynamics of IPD, as at the time of writing that provides the most complete data set to inform the serotype dynamics to date in England.

Our population structure for England, which captures the continued growth of the elderly population, is informed by historic census estimates [24] and projections [25] by the Office of National Statistics.

The uptake of PCV vaccine in children has been relatively high, with 93% completing PCV7 vaccination (from 2006 to 2010), and 91% completing PCV13 vaccination (from 2010 to 2023) [26, 27]. We assume that uptake will continue at 91% in our future projections. For older individuals, PPV23 was available to the over 80s from August 2003, to the over 75s from August 2004 and to the over 65s since April 2005. Coverage of PPV23 in those 65 years of age or older has been consistently between 69% and 71% from 2006 to 2021 [28]; more recent data has shown a slight rise to 73% [29]. In our future projections, we therefore assume 72% coverage for elderly vaccination with either PPV23 or PCV20.

Vaccine effectiveness estimates are complex, as there will be different effectiveness estimates against disease (pCAP and IPD) and against carriage. We assume the same estimates against both IPD and pCAP for PCVs. Data from England and Wales suggests that efficacy against disease in full vaccinated children from PCV7 is 93% Andrews *et al.* [30], while efficacy from PCV13 (aside from serotype 3) is around 90%. We also expect the protection provided by vaccination to decline over time. We infer this decline in immune protection over time provided by vaccination. PPV23 has been shown to be ineffective against carriage and pCAP [31]; against IPD its efficacy drops from 41% within two years of vaccination to 23% for those vaccinated more than 5 years ago [32].

Vaccine efficacy against carriage is substantially less than that against disease. A meta-analysis of ten studies found that efficacy against carriage was estimated to be 57% for PCV7 [33]. For PCV13 studies in Malawi and Vietnam estimate efficacy against carriage between 62-65% for fully vaccinated individuals [34, 35]. However, a model inference using English data suggested that the efficacy of PCV13 was far lower [13]. Given this uncertainty, we incorporate the efficacy of PCV13 against carriage into our parameter inference scheme.

Epidemiological data on carriage, pCAP and IPD come from a variety of sources. Age-stratified data on serotype carriage with age are provided by studies that swab members of households with young children either longitudinally (in 2001/2002) [21] or single samples (in 2008, 2012, 2015 and 2018) [22]. Data are available on approximately 2400 individuals, although the sampling methodology means strong bias towards younger age-groups. We note that serotypes 1 and 5 were not detected in these carriage studies but are known to cause disease.

While many studies have focused on CAP in general, very few have considered the causative agent. Detailed data from 2008 to 2023 from Nottingham Hospitals sheds light on the distribution of serotypes [36] and the age-distribution in adults [37]. The risk of pCAP in children aged 15 or under is based on a reported incidence rate of CAP estimated to be 144 in 100,000 [38], of which 46% (66 per 100, 000 are due to pneumococcal infection [39]. We further assumed the age-distribution of pCAP in children to have the same profile as IPD (which we judge as a reasonable assumption given the similarity in age-distributions for adults).

The UK Health Security Agency (UKHSA) and before that Public Health England (PHE) monitors IPD cases in hospitals. In each calender year, cases are stratified into six age-groups (under 2, 2-4, 5-14, 15-44, 45-64 and over 65 years old), serotype (where available, with over 90% having their serotype determined since 2010) and the total number of IPD cases. These IPD data contains over 114,000 serotyped cases between 2000 and 2023.

Before the introduction of the PCV7 vaccine in England in 2006, infants (under 2 years of age) were most at risk from IPD (Fig. 1(a)), experiencing over 50 cases per 100,000; although due to the greater population size IPD was most common in those over 65 years old (over 46% of cases are in the oldest age-group). These early cases of IPD were caused by serotypes (especially serotype 14, which caused 18% of all serotyped cases) that were subsequently targeted by the PCV7 vaccine (Fig. 1(b)). With the introduction of infant pneumococcal vaccination programmes using PCV7 starting in 2006 and then PCV13 in 2010 (denoted by arrows in Fig. 1(a)), the risk of infant IPD sharply decreased to typically less than 15 IPD cases per 100,000 per year, dominated by serotypes that are not within the PCV13 vaccine. Since 2010, IPD rates for PCV7 serotypes, and to a lesser extent those of PCV13, have then remained low, including among the elderly (Figs. 1(b) and 1(c)) through indirect protection. However, there has also been a pattern of resurgence of the non-vaccine serotype IPDs (that are not part of PCV13) due to serotype replacement, especially among the elderly (Fig. 1(d), Fig. 1(e)). In 2023 the rate of IPD cases in the over 65s was 1.3, 5.9, 8.0 and 10.4 per 100,000 for the PCV7 serotypes, the additional six serotypes in PCV13, the additional seven serotypes in PCV20 and non-vaccine serotypes, respectively.

As a consequence, IPD is now predominately a disease of the elderly; the IPD case rate per 100,000 per year (across all serotypes) in those aged 65 years and above has remained relatively high since 2000 (frequently above 20 IPD cases per 100,000 per year). The only change to this pattern was during 2020 and 2021 when containment measures associated with the SAR-CoV-2 pandemic also reduced the transmission of pneumococcal infection. We hypothesise that the burden IPD places on the health services is only likely to increase without further measures, due to the projected increase in size of the population aged over 65.

### 2.2 Overview of the pneumococcal transmission model

Our pneumococcal transmission model is a deterministic system of ordinary differential equations that recognise serotype and age-structure. Unlike traditional compartmental models that seek to place individuals into discrete non-overlapping compartments (e.g. a compartment for individuals infected with serotype 6A, resistant to serotypes 1,3 and 22F, but susceptible to all other serotypes), here we adapt the lower-dimensional framework of Andreasen *et al.* [19]. In particular we model the number of individuals that are susceptible, vaccinated, infected (which we term as having ‘carriage’ status) or resistant to each serotype - ignoring their status with respect to other serotypes (Fig. 2). This vastly reduces the computational overheads with simulating the dynamics, although it does complicate the model formulation for serotype interactions.

**Fig. 2.**
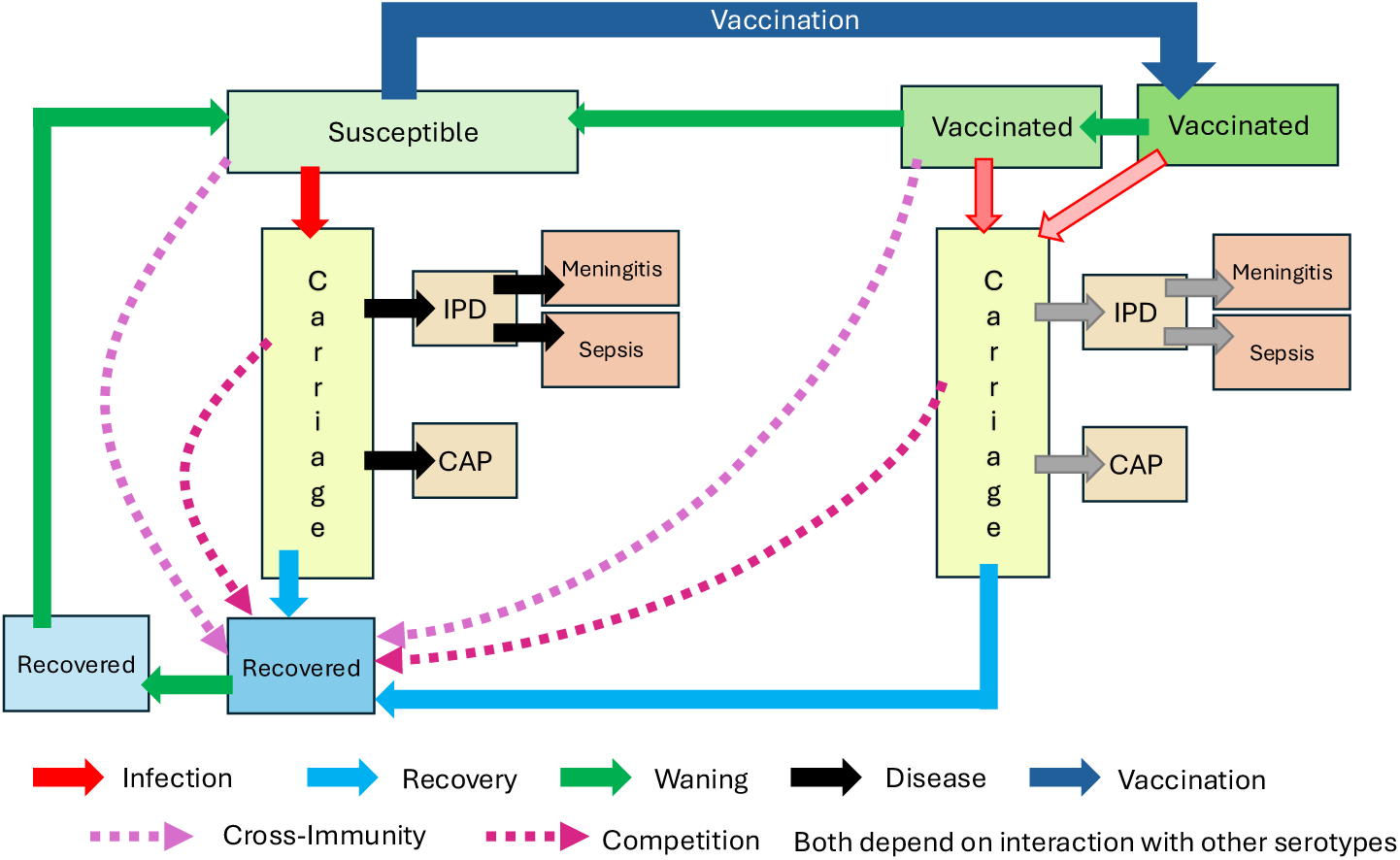
Schematic of the MEMVIE pneumococcal disease model for a given serotype. For a given serotype we track how many people are susceptible, vaccinated, infected (which we term as having a ‘carriage’ status) or resistant to that serotype. Those with carriage have an age- and serotype-dependent probability of developing community-acquired pneumonia (CAP) and/or invasive pneumococcal disease (IPD). We can consider conditions arising from IPD, such as meningitis and sepsis. The compartments are replicated for the three types of pneumococcal conjugate vaccine (PCV7, PCV13, PCV20; whereas PPV23 only acts on the rate of IPD development), nine age groups (0 up to 3 months, 3 up to 12 months, 12 months up to 2 years, 2-4 years, 5-14 years, 15-44 years, 45-64 years, 65-75 years, 75+ years) and 26 serotypes. The model includes interactions between serotypes in the form of cross-immunity (if infected by one serotype that could result in a protective effective towards another serotype) and competition (replacement of carriage by a different serotype).

Disease (IPD and pCAP) occurrence is proportional to the carriage of each serotype and age-group; with the constant of proportionality being the product of an age-dependent factor and a serotype dependent factor. To account for vaccine efficacy with respect to disease, infected individuals are separated by their vaccination status. PPV23 vaccination only affects the development of IPD, so is irrelevant to the infection dynamics and is incorporated as part of the disease process. The epidemiological transmission dynamics are unaffected by disease.

The 26 serotypes considered in the model were: those contained in the PCV7 vaccine (4, 6B, 9V, 14, 18C, 19F, and 23F); additional serotypes in the PCV13 vaccine (1, 3, 5, 6A, 7F and 19A); additional serotypes in the PCV20 vaccine (8, 10A, 11A, 12F, 15B, 22F and 33F); and serotypes not contained in the PCV vaccines (2, 6C, 9N, 17F, 20 and all Others). There are two processes that govern the interaction between serotypes - otherwise the serotype dynamics would be independent. These are: competition, which occurs when the serotype being carried is replaced by infection with a new serotype; and cross-immunity when infection with one serotype confers partial immunity to other serotypes (Fig. 2)

In addition to stratifying the population by serotype infection status, we stratified the population into nine age groups: 0 up to 3 months, 3 up to 12 months, 12 months up to 2 years, 2-4 years, 5-14 years, 15-44 years, 45-64 years, 65-75 years, 75+ years. The partitioning of the youngest three age groups (0 up to 3 months, 3 up to 12 months, 12 months up to 2 years) was a pragmatic choice to enable allocation of vaccination as individuals transition between those age-groups.

We provide expanded model details, including the underlying equations, in Section S2.

We fit the model to the available carriage and IPD data in England using a Bayesian, likelihood-informed approach. We accounted for the reduced level of serotyping over the first ten years. We note that given the disparity of size in the two data sources (carriage 2400 individuals, IPD 114,000 serotyped cases), the IPD data provides a greater constraint on the parameters. The main parameters inferred are: serotype-specific transmission, recovery and fitness; age-dependent and serotype-dependent case-carriage ratios for both IPD and pCAP; duration of vaccine protection and efficacy of the PCV13 vaccine against carriage; and the impact of COVID-19 restrictions on transmission. Although our model is based on nine age-groups, we aggregate our projected IPD cases to match the six age-groups in the UKHSA data (*<* 2, 2-4, 5-14, 15-44, 45-64, and 65+) The Supporting Information provides mathematical details of fitting the model output to the available data on carriage (Section S4), IPD (Section S5) and pCAP (Section S6).

### 2.3 Overview of the health economic model

The health economic calculations required monetary costs and a measure of disease impact on health (captured through the loss of Quality-Adjusted Life Years, QALYs) associated with each level of disease outcome: IPD, pCAP, post-infection sequela and death. We applied an annual discounting rate of 3.5% to costs (both health care and vaccination costs) and health effects.

We obtained monthly hospitalisation data from the Hospital Episode Statistics (HES) database through the Department of Health. We identified cases by the presence of the relevant diagnostic discharge codes listed in the patient’s discharge record. Each observation comprised a Finished Consultant Episode (FCE), which measures the time the patient spends under the care of a particular consultant. By mapping Healthcare Resource Group (HRG4) codes associated with diagnoses to reimbursement costs, we acquired hospitalisation costs per IPD and pCAP (Table 1). A detailed description of this cost estimation procedure is provided in the Supporting Information (see Section S7).

**Table 1.** Health episode costs and QALY losses used in the health economic assessment. The Combined Health Economic Costs are calculated at £20,000 per QALY. QALY losses for a hospitalisation episode (without sequelea) are between 0.0015 and 0.025 (between £30 and £500). These are included in the Combined HE Costs, but not shown in the table for brevity. We report all values to at least 2 s.f.

| Age<br>(years) | Costs per<br>hospitali-<br>sation<br>episode<br>(£) |  | Risk of<br>mortality |  | QALY<br>loss for<br>mortal-<br>ity<br>(both) | Mening-<br>itis risk<br>(IPD) | QALY<br>loss<br>from<br>Menin-<br>gitis<br>(IPD) | Combined<br>HE Cost<br>(£) |  |
| --- | --- | --- | --- | --- | --- | --- | --- | --- | --- |
|  | IPD | pCAP | IPD | pCAP |  |  |  | IPD | pCAP |
| < 2 | 4164 | 2339 | 2.8% | 0.7% | 23.6 | 12.0% | 5.69 | 31,260 | 5691 |
| 2-4 | 4292 | 2456 | 2.8% | 0.7% | 23.5 | 5.6% | 5.66 | 23,869 | 5771 |
| 5-14 | 3939 | 2268 | 9.4% | 0.7% | 22.8 | 3.8% | 5.49 | 51,230 | 5499 |
| 15-44 | 5483 | 1516 | 9.4% | 0.7% | 20.2 | 2.2% | 4.87 | 45,866 | 4377 |
| 45-64 | 6187 | 1654 | 9.4% | 1.8% | 14.8 | 2.8% | 3.57 | 36,237 | 7097 |
| 65-74 | 6276 | 1815 | 24% | 7.6% | 9.8 | 1.3% | 2.36 | 54,170 | 15,010 |
| >75 | 6276 | 1812 | 24% | 12.1% | 3.9 | 1.3% | 0.98 | 25,884 | 11,534 |

Determining the impact of disease in terms of QALY losses is divided into three separate calculations: age-dependent losses due to hospitalisation (assuming no long-lasting disease); age-dependent losses due to mortality; and losses due to long-lasting sequelae, which combines the age-dependent risk of meningitis with the risk of multiple sequelae and the associated reduction in QALYs.

For QALY loss per hospitalisation, we sourced health state utility values from the literature. Due to limited evidence on the health-related quality of life (HRQoL) for individuals hospitalised with septicaemia, we collectively applied to IPD the utility value for meningitis hospitalisation [40]. The utility values for pCAP by age group were available only for adults aged 19 and above; therefore, we assumed that individuals under 19 had the same utility value as those aged 19–35 years [41]. QALY losses per hospitalisation are shown in (Table 1).

To inform QALY losses due to mortality, we took year-of-age and sex-stratified estimates of quality adjusted life expectancy for the population of England from McNamara *et al.* [42], with values discounted at a 3.5% annual rate. To obtain our estimated QALY loss associated with mortality for each age group (as stated in Table 1), we weighted age and sex specific estimates for quality adjusted life expectancy from McNamara *et al.* [42] by the population structure for England recorded in the 2021 census [24].

Finally, we sourced QALY losses for individuals living with deafness, mild hearing loss, epilepsy, mild mental retardation, severe retardation and tetraplegia, and leg paresis from the literature and reported on a per year basis [43] (Table S2). We combined these QALY loss estimates for each sequelae with the risk of each sequela and the life-time QALY (accounting for discounting). Together, these produced a final measure of the total expected QALY losses for each age-group due to meningitis (Table 1).

### 2.4 Calculation of willingness to pay (WTP) thresholds

We used our pneumococcal transmission model to project the infection dynamics forwards in time for the next 50 years; the infection data together with our (age- and serotype-specific) estimates of case-carrier ratios allows us to estimate the number of IPD and pCAP each calendar year in each age group. By comparing the total costs and QALY losses between two simulations with different vaccination strategies, we can calculate the willingness to pay (WTP) threshold for each vaccine dose. In particular, we can calculate the WTP for a given vaccine programme compared to no vaccination (assuming an administration price of £10.06 per dose in children and adults, rising to £12.06 for children in 2026) or calculate the additional WTP for one vaccine compared to another (e.g., the additional WTP for a single dose of PCV20 above a single dose of PCV13). In the latter case, the administration price is irrelevant. Throughout, we followed the guidelines established by the Joint Committee on Vaccination and Immunisation (JCVI - an expert scientific advisory committee that advises the UK government on vaccination and immunisation matters) working group on Uncertainty in Vaccine Evaluation and Procurement (see Annex 5, Section 3 of the JCVI code of practice [23]). We calculated two quantities: a central estimate of the WTP threshold (using the maximum likelihood parameter estimates and valuing one QALY at £20,000) and the WTP threshold for which 90% of scenarios were cost-effective (using £30,000 per QALY and fully accounting for parameter uncertainty). The reported WTP is the minimum of these two quantities.

We evaluated all vaccine programme change scenarios over four time horizons: 5, 10, 25 and 50 years; implemented by truncating the simulations. Short time horizons provide insight into the likely dynamics without serotype replacement. However, short time horizons are also necessarily biased as although they include the full cost of vaccination for the period, the impact of protection may last beyond the time horizon; this bias in non-captured protective effects beyond the time horizon is diminished in longer simulations due to discounting. As we wish to calculate the long-term impact of amendments to the pneumococcal vaccination programme, even if there may be other changes to the programme in the future, we therefore believe the 50-year time horizon to be the most appropriate.

### 2.5 Model simulation of retrospective and prospective scenarios

Ed says: “Suggesting header and text edits to align methods content grouping with the simulation results subsections in the overall results section of the manuscript.”

We perform three sets of (retrospective or prospective) simulations considering changes to the vaccine schedule at three distinct time points (2026, 2010 and 2026). We compute WTP thresholds by the comparison of different scenarios (either at the maximum likelihood parameters or accounting for uncertainty, and for the four time horizons: 5, 10, 25 and 50 years).

#### Changes to the paediatric and adult pneumococcal vaccination programme in 2026

We first considered scenarios involving a potential switch in vaccine product in England from 2026 onwards in the paediatric vaccine programme, from PCV13 to PCV20, and/or the adult pneumococcal vaccine programme, from PPV23 to PCV20. These provide WTP thresholds for decisions that are facing the UK vaccine policy-makers.

#### Changes to the paediatric pneumococcal vaccination programme in 2006, 2010 and 2026

We next ran a retrospective analysis at historical decision points, as well considering changes in 2026. In 2006, the options are either adopt the PCV7 vaccine in infants or not; in 2010 the options introduce the PCV13 vaccine, continue with the the PCV7 vaccine or stop paediatric vaccination; while in 2026 we can vaccinate with PCV20, PCV13 or PCV7, or not vaccinate. By comparing pairs of strategies, we evaluate the WTP at each of the three decision points (either at the maximum likelihood parameters or accounting for uncertainty, and for the four time horizons). For all simulations, we either assume continuation of PPV23 vaccination in adults aged 65 or over, or a switch in 2026 from PPV23 to PCV20.

#### Willingness to pay threshold sensitivity to PCV20 efficacy

Vaccine efficacy is an essential factor that determines the impact of any programme and hence can substantially alter the WTP per vaccine dose. Lower efficacy, against carriage of pneumococcal diseases or against disease (IPD and pCAP), would generally be anticipated to result in a lower WTP for the vaccine. There is some intuition that higher valency vaccines could have lower efficacy. Nonetheless, throughout we have made the parsimonious assumption that the efficacy of PCV20 (against disease and carriage) matches the efficacy of PCV13 (90% against disease, and an inferred value of around 50% against carriage).

Given the anticipated sensitivity of WTP per vaccine dose to vaccine efficacy, for our final analysis we computed WTP thresholds for a dose of PCV20 over PCV13 for different PCV20 efficacy values. We considered efficacy against disease and carriage separately. We made the intuitive assumption that the efficacy against disease should always be greater than efficacy against carriage. We varied efficacy against carriage from 0.4 to 0.75 (in increments of 0.05) and varied efficacy against disease from 0.6 to 0.925 (in increments of 0.025). We made these WTP calculations assuming the change from PCV13 to PCV20 in the paediatric pneumococcal vaccination programme begins in 2026. As previously, we produced WTP threshold measures for the four different time horizons of 5 years, 10 years, 25 years and 50 years.

## 3 Results

### 3.1 Fitting the pneumococcal transmission model to historical data

Using the inference procedure described in Section 2.2 and in the Supporting Information (Sections S4 to S6), we fit our transmission model to observed IPD and carriage data to maximise the likelihood associated with each of the 26 serotypes under consideration. Given the abundance of IPD data compared to that from carriage studies, capturing the disease dynamics over the 24 years contributes significantly more to the likelihood than the carriage (infection) dynamics.

For most serotypes there was a good match between the data and the model results for age- and serotype-specific IPD (Fig. 3; agreement with the carriage data is shown in Fig. S4. We readily capture the decline in vaccine serotypes following the introduction of PCV7 in 2006 (serotypes 4, 6B, 9V, 14, 18C, 19F, 23F) and PCV13 in 2010 (which additionally included serotypes 1, 3, 5, 6A, 7F and 19A). We also have reasonable correspondence for the rise in non-vaccine serotypes. Unsurprisingly, given the complexities of the underlying processes, there are some discrepancies between the model and the data. The most notable discrepancy is we underestimate the rise in ‘Others’ group, which contains all serotypes excluding the 25 individual ones considered in the model. We postulate that this discrepancy is because the true dynamics are composed of the rise, fall and competition between multiple serotypes that cannot be effectively captured by this aggregate. However, given these are non-vaccine serotypes (NVT), we do not expect this misfit to adversely affect the willingness to pay for PCV vaccines (a more detailed comparison for a few serotypes is presented in Fig. S5). Another discrepancy, which highlights the complexity of IPD dynamics, is that while all other serotypes are dominated by disease in the oldest age-group, for serotype 1 the greatest number of disease cases is in the 16-44 year olds. Given we make the parsimonious assumption that case-carrier ratios are a product of serotype and age-group specific values, it follows there is an inability of the model to capture the anomalous age-distribution of IPD caused by serotype 1.

**Fig. 3.**
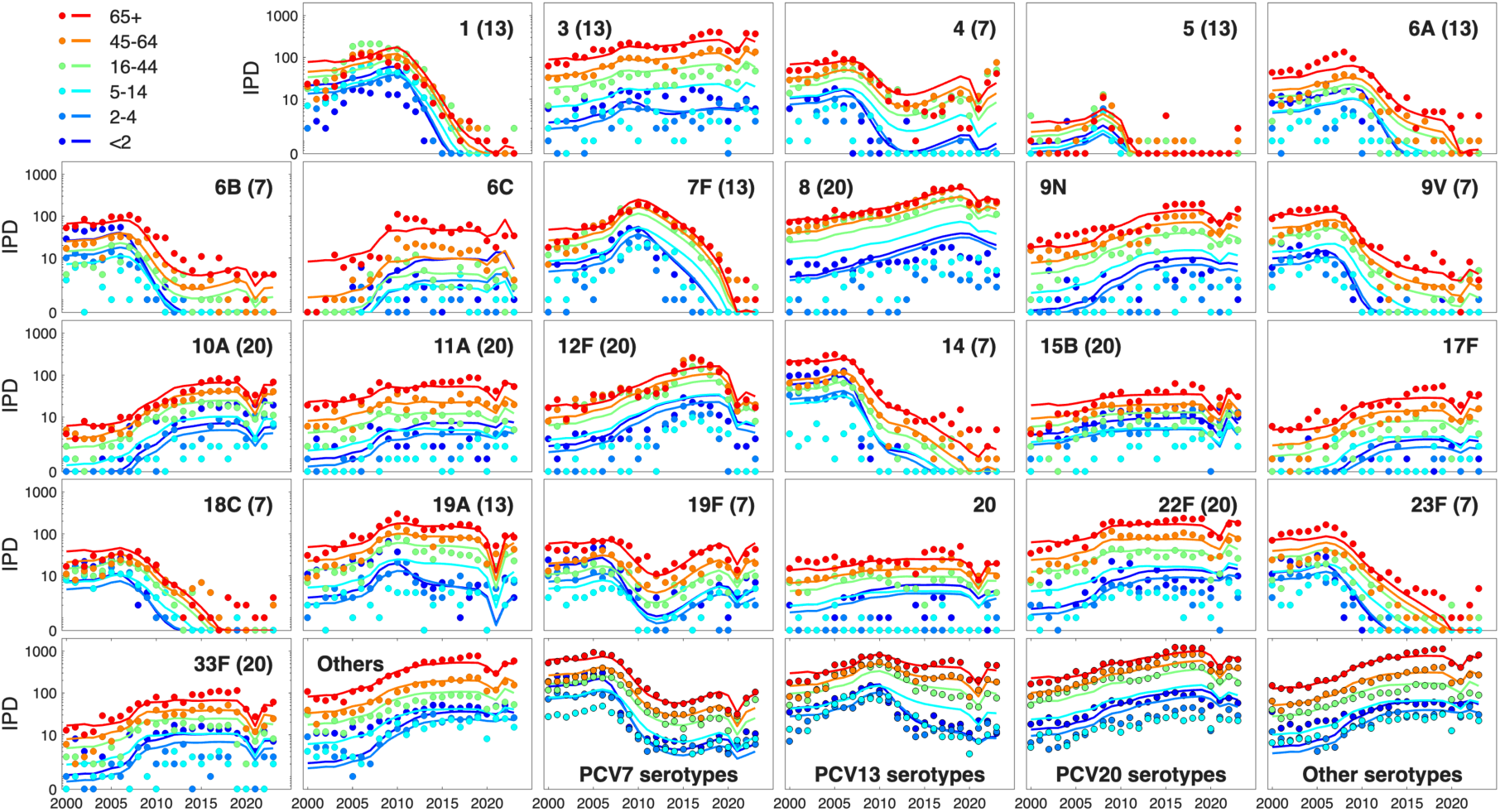
Match between best fit model results (lines) and data (dots) for age and serotype specific invasive pneumococcal disease (IPD). Colours denote the six different age groups. Serotype 2, which remains low throughout is not shown. IPD cases were a Poisson sample from infected individuals based on the case-carrier ratios. The last four plots show the sum of serotypes in PCV7, PCV13 (but not PCV7), PCV20 (but not PCV13) and all other (non-vaccine) serotypes. The results are plotted on a logarithmic scale to better display the range of values.

Although the introduction of PCV7 in 2006 led to a decline (though not elimination) of IPD due to PCV7 serotypes for all age groups, especially among infants, there was a later rise in IPD due to the remaining serotypes (Fig. 1(a) and Fig. 3). In our model, these trends are driven by serotype replacement, where suppression of some pneumococcal serotypes allows for the emergence of others. A similar pattern was observed for the six additional serotypes included in PCV13; its introduction in 2010 was accompanied by a resurgence of non-PCV13 serotypes (Fig. 1(a) and Fig. 3). We note that the observed decline in IPD caused by PCV7 serotypes after the introduction of PCV7 in 2006 is larger than the decline in PCV13 serotypes when PCV13 was introduced in 2010. We attribute this difference to the limited effectiveness of the PCV13 vaccine against serotype 3 [44], which remains a leading cause of IPD throughout the time course investigated.

Lastly, we note that there are distinct differences between the pre- and post COVID-19 pandemic period, driven by the drop in transmission during 2020 and 2021. While the five serotypes with the highest IPD have remained consistent (‘Others’, 3, 8, 9N and 22F), post-pandemic we have observed fewer cases of IPD (suggesting a slow dynamic recovery) with serotype 8 being particularly impacted and ‘Other’ serotypes dominating.

### 3.2 Willingness to pay thresholds for changes to the paediatric and adult pneumococcal vaccination programme in 2026

We begin by considering the historic costs of pneumococcal disease comprising direct NHS costs and QALY losses (at £20,000 per QALY), all based on current costs to remove the effects of inflation (Fig. 4). The majority of costs are associated to QALY losses due to IPD or pCAP. Over the three time periods (before vaccination in 2000-2005, during PCV7 vaccination in 2007-2009 and during PCV13 vaccination in 2011-2019), there is an overall slight reduction in the total costs (from £474M to £391M per annum based on current prices), a notable decline in the costs associated with the younger age-groups, but a relative increase in the QALY losses with older age groups dominating.

**Fig. 4.**
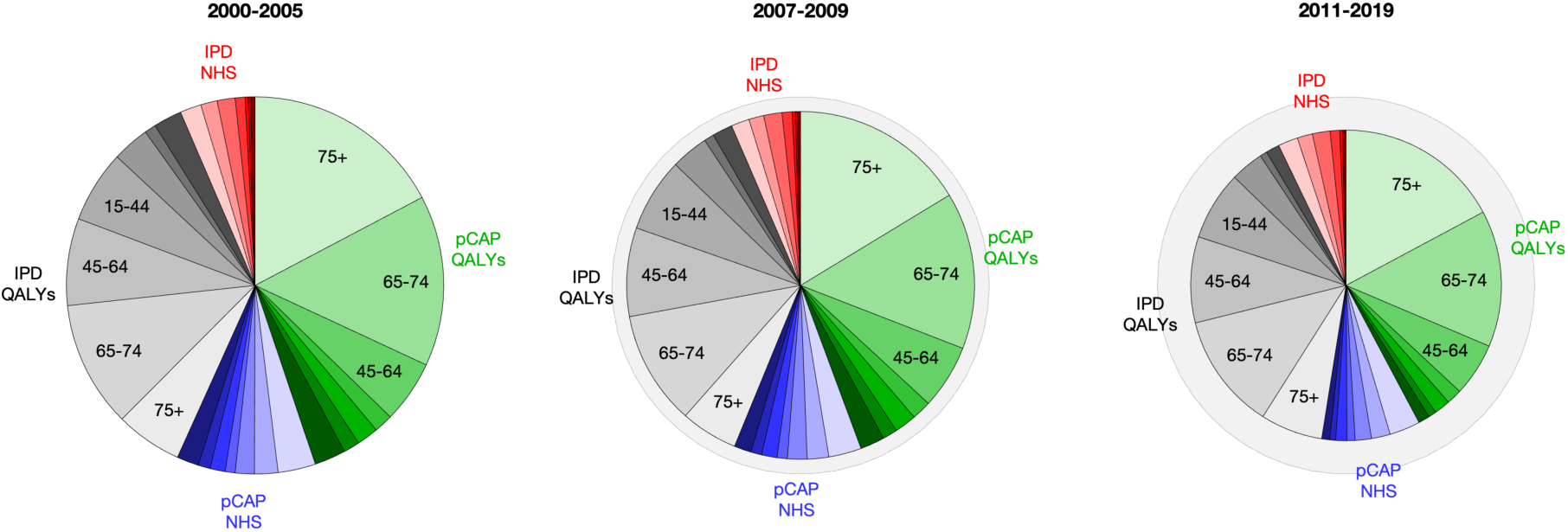
Retrospective analysis of the annual costs of pneumococcal disease. We consider three separate time-scales: before vaccination (2000-2005), during PCV7 vaccination (2007-2009) and during PCV13 vaccination (2011-2019). Pie charts show the average yearly costs for QALY loss due to IPD (grey at £20,000 per QALY), NHS costs of treating IPD (red), QALY loss due to pCAP (green at £20,000 per QALY) and NHS costs of treating CAP (blue); age groups are shaded from dark (youngest) to light (oldest). The size of the pie charts is relative to the overall costs (£474M, £437M, £391M). The outer dashed circle highlights the reduction from the pre-vaccination scenarios.

From these base-line costs, we compute WTP thresholds for scenarios involving a potential switch in vaccine product from 2026 onwards in the paediatric and/or adult pneumococcal vaccine programmes in England (Fig. 5). For all but one of these scenarios, we found the central estimate of the WTP threshold (using the maximum likelihood parameter estimates and valuing one QALY at £20,000) was less than the 90% prediction interval for the WTP threshold using £30,000 per QALY, and therefore remains our quantity of interest. The exception is for the longer time-horizon WTP thresholds due to a switch from PPV23 to PCV20 in the elderly while keeping PCV20 among the infants.

**Fig. 5.**
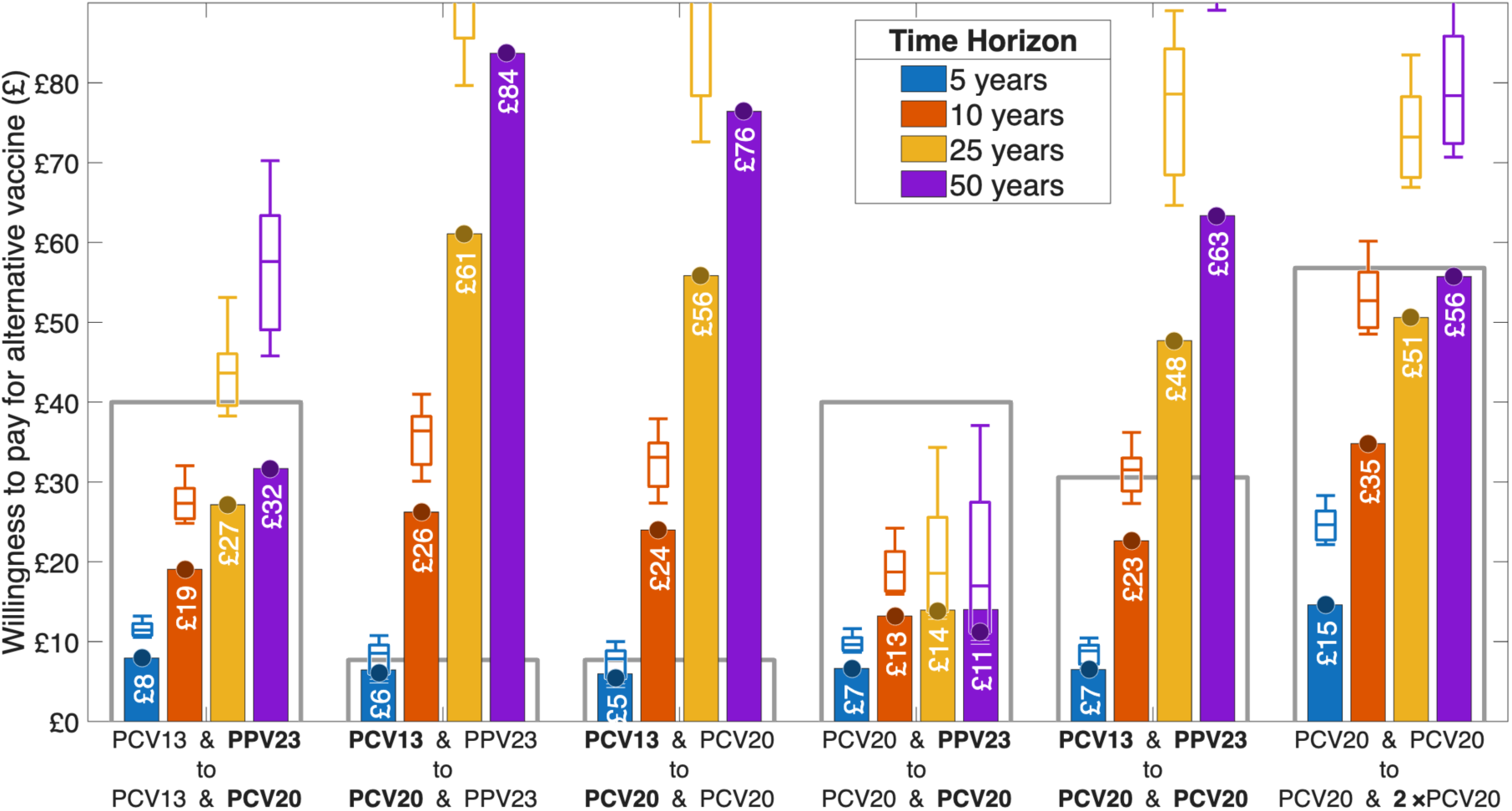
Willingness to pay thresholds across four different time horizons (5, 10, 25 and 50 years) for changes in paediatric and adult vaccination. WTP thresholds for changes in 2026, either switching from PCV13 to PCV20 in infants and/or switching from PPV23 to PCV20 in older adults; the switch is shown in bold. The final set of results correspond to additional vaccination of 75 year olds using PCV20, and includes the additional administration fee. The bars correspond to the central estimate of the WTP threshold (using the maximum likelihood parameter estimates and valuing one QALY at £20,000), while the error bars correspond to the prediction interval for the WTP threshold using £30,000 per QALY and accounting for parameter uncertainty. The dots correspond to the minimum of these two quantities and therefore the WPT under the JCVI uncertainty criterion. The open grey boxes correspond to the cost of the switch using vaccine list prices of £16.80 for PPV23, £49.10 for PCV13 and £56.80 for PCV20.

The immediate conclusions are that a switch to PCV20 (from PCV13 in infants or PPV23 in elderly) is cost-effective if the vaccine can be procured for a sufficiently low price. The willingness to pay generally increases as longer time horizons are considered and the timescales of vaccination and vaccine action become similar. There is a consistently a greater willingness to pay for PCV20 replacing PCV13 in infants (second and third scenarios in Fig. 5), compared to PCV20 replacing PPV23 in the elderly (first and fourth scenarios in Fig. 5). There is also a small amount of cross-reaction, such that PCV20 in infants is less cost-effective if PCV20 is also deployed in the elderly (comparing second and third scenarios); and similarly PCV20 in the elderly is less cost-effective if it is also deployed in infants (comparing first and fourth scenarios). Over a 50-year time horizon, the willingness to pay for each dose of PCV20 in infants (above the price paid for PCV13) is £76 or £84, depending on the vaccine used in older adults. Over the same time horizon, the willingness to pay for PCV20 (over PPV23) in adults is £11 or £32, depending on the vaccine deployed in children. When considering the combined switch from PCV13 and PPV23, to PCV20 vaccination in both infants and adults, the willingness to pay is £63.

Given that pneumococcal disease is now predominantly in elderly individuals, and given that protection from vaccine wanes over time, it is prudent to consider a later ‘booster’ vaccination strategy. The last set of bars in Fig. 5 corresponds to the willingness to pay for an additional dose of PCV20 given at age 75. In keeping with vaccination at age 65, we assume an uptake of 72% and make the simplifying assumption that uptake is random (in both those that did and did not get vaccinated at age 65). The willingness to pay threshold is £56 (over a 50 year time horizon) even when administration costs (of £10.06) are considered. Unlike the other willingness to pay values that are related to the cost of PCV20 above the cost of existing vaccines, as this is an addition to the schedule, the threshold of £56 relates to the true cost per dose.

It is important to set these willingness to pay thresholds in relation to the vaccine list prices (Fig. 5 grey bars). PPV23 is the least expensive vaccine (£16.80) while PCV13 and PCV20 are considerably more expensive (£49.10 and £56.80 respectively); therefore, a switch from PCV13 to PCV20 only needs to be associated with a relatively small willingness to pay, whereas a switch from PPV23 to PCV20 needs to deliver far more health benefits to be cost effective. As such, we find a switch from PCV13 to PCV20 paediatric vaccination is cost effective (at the list prices) given time horizons of 10 years or more. Whereas, the switch from PPV23 to PCV20 in the elderly is not cost effective at the list price. The addition of an extra dose of PCV20 at age 75 could be cost effective over a 50 year time horizon if it could be purchased just below the list price.

### 3.3 Willingness to pay thresholds for changes to the paediatric pneumococcal vaccination programme in 2006, 2010 and 2026

We expand upon our previous results by including a retrospective analysis at historical decision points as well as changes in 2026 (Fig. 6(a)). In particular, we consider the dynamics (total number of IPD cases) and WTP thresholds at three change points: 2006 when PCV7 was introduced, in 2010 (when PCV13 was introduced), and 2026 (when PCV20 may be introduced). At each point, we consider the WTP and dynamics of all viable alternatives based on the number of available vaccines; as such in 2007 the decision is between no vaccination and PCV7, in 2010 the decision is between keeping with PCV7, switching to PCV13 or stopping vaccination, while in 2026 the options are between the three vaccines PCV7, PCV13 and PCV20 or stopping vaccination. In each case, we calculate the WTP threshold of future options against the vaccine programme up to that time. Again, we generated WTP threshold estimates for time horizons of 5, 10, 25 and 50 years, respectively, providing some understanding of the longer-term roles of serotype replacement. In all cases (where the WTP is positive), the JCVI uncertainty criteria is determined by the maximum likelihood parameters at £20,000; hence, for clarity, this is all that is shown.

**Fig. 6.**
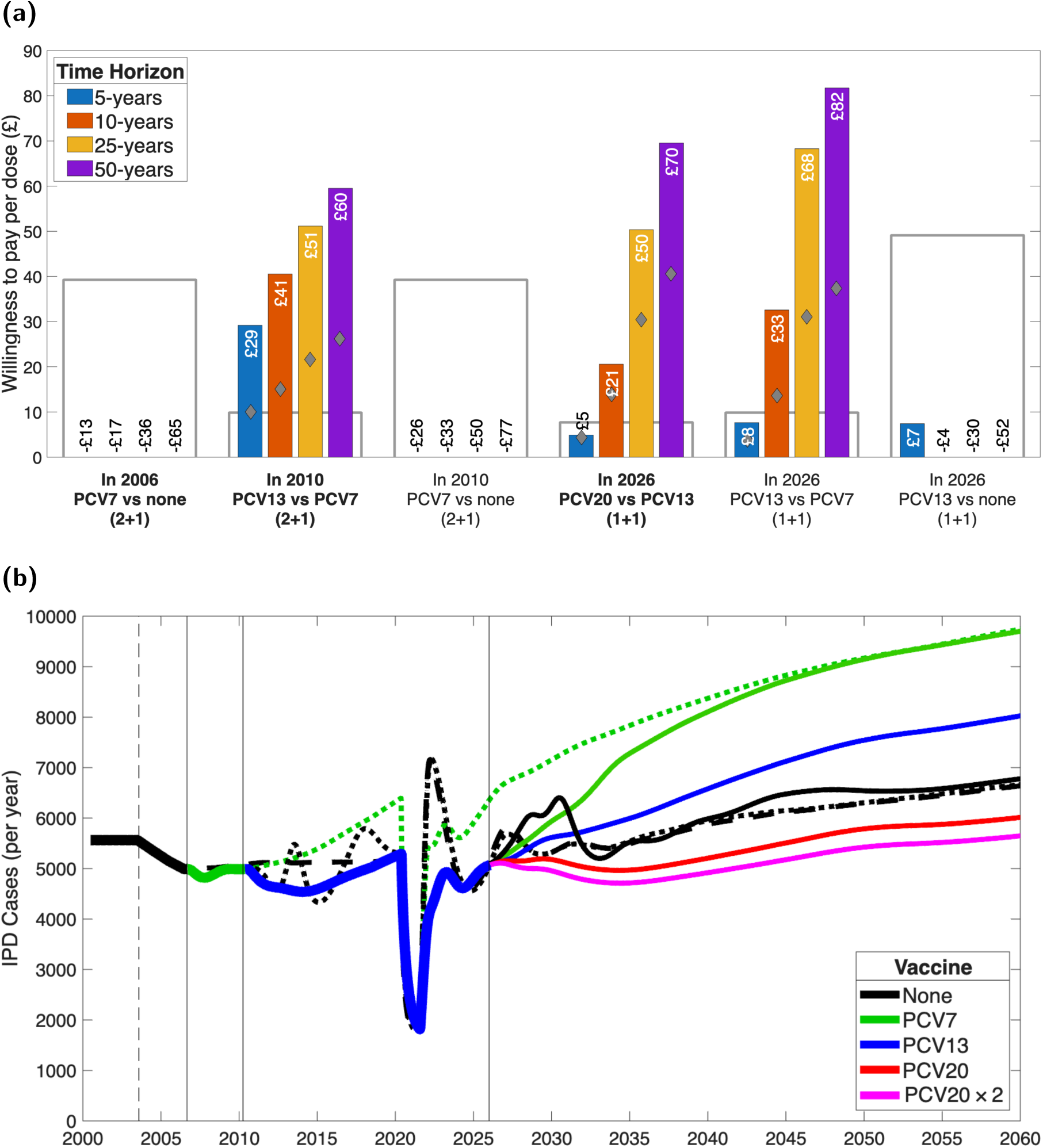
Willingness to pay thresholds and associated changes in IPD for changes in vaccination in 2006, 2010 and 2026. **(a)** WTP of potential paediatric vaccine changes at £20,000 per QALY (this is consistantly the minimum of the uncertainty criterion). Diamonds show the value for IPD costs only; the open grey bars correspond to the cost of the switch using vaccine list prices of £39.25 for PCV7, £49.10 for PCV13 and £56.80 for PCV20. Vaccine switches before 2020 are assumed to adopt a (2+1) vaccination scheme. **(b)** Estimated IPD for the different strategies considered. Comparisons reflect the availability of vaccines at the time: in 2006 there was a choice between introducing PCV7 (solid green) or not vaccinating (dashed black); in 2010 the choice was to switch to PCV13 (solid blue), remain with PCV7 (dotted green) or stop vaccination (dotted black); while in 2026 there are four choices (thinner lines), a switch to PCV20, remaining with PCV13, a switch to PCV7 or stopping vaccination. The magneta line shows additional vaccination with PCV20 for those aged 75 starting in 2026. (Throughout we start PPV23 vaccination in older adults in 2003 and switch to PCV20 in 2026.)

In 2006, the short-term WTP threshold for PCV7 (compared to no-vaccination, and therefore including the administration cost of £10.06 per dose) is negative: -£17 if PCV7 is administered with a 2+1 schedule, for a ten-year time horizon. At longer timescales, the cost-effectiveness reduces even further as there is substantial serotype replacement and the total number of IPD cases increases (thick green line and dotted green line in Fig. 6b). Unsurprisingly, by 2010 with the rise of non-PCV7 serotypes, there is a substantial WTP threshold for PCV13 over the cost paid for PCV7, and far fewer IPD cases (comparing thick blue line to dotted green line from 2010 onwards, Fig. 6b). This WTP threshold for PCV13 (against PCV7) peaks at £60 over a 50-year time horizon; but even over a 5-year time horizon the additional WTP threshold is £29. At the same point, continuing with PCV7 has a negative WTP compared to stopping vaccination for all time horizons.

These negative WTP values for PCV7 are in stark contrast to the initial modelling results [11], which suggested PCV7 would be cost-effective at around £10-15 per dose. We explain this discrepancy as being driven by serotype replacement, which could not have been predicted at the time PCV7 was introduced. Although PCV7 vaccination caused a notable decline in IPD cases in the youngest age groups (Fig. 1(a)) it did not prevent the expansion of disease in the oldest ages. A simple statistical fit to the serotype IPD data generates similar projections for the willingness to pay threshold (Section S8).

In 2026, along with the assumed start of PCV20 vaccination in the elderly, there are four potential decisions about future vaccination. A switch to PCV20 (from PCV13) has a WTP threshold of £76 (as was observed in Fig. 5); this is to be expected given the lower number of IPD cases with this scenario (red line Fig. 6b). At this time point, there is a significant WTP threshold for keeping with PCV13 rather than switching to PCV7; although the WTP for maintaining PCV13 vaccination compared to stopping vaccination substantially diminishes over time and becomes negative at a 10-year time horizon. These results are broadly attributable to serotype replacement and the case-carrier ratio for the strains that are present. In the long-term, we estimate that, relative to the no-vaccination model, PCV20 leads to a suppression of IPD cases by 9%, while PCV13 and PCV7 lead to increases of 22% and 46% respectively.

Finally, Fig. 6b also shows the impact of PCV20 in infants combined with two doses of PCV20 in the elderly at age 65 and 75 (pink line), which leads to a further decline in IPD. Given that this decline is predominantly in the elderly who suffer the largest health burdens associated with pneumococcal infection (Fig. S6 and S7), the health economic impact associated with this decline is considerable (hence the high WTP thresholds in Fig. 5).

### 3.4 WTP sensitivity to PCV20 efficacy

The data on IPD and carriage for England has allowed us to estimate an efficacy against carriage for the PCV13 vaccine of 50% (pink dot), compared to 57% for PCV7 (cyan dot) [33]. However, epidemiological data on the real-world impact of PCV20 is lacking. Most data on PCV20 efficacy has been derived from antibody studies [45]. Antibody studies are difficult to correlate quantitatively to protection [46], although there is some evidence of increased break-through infections with higher valency vaccines [47]. Throughout the paper, we have assumed that the protection offered by PCV20 is the same as that of PCV13 (50% against carriage and 90% against disease), but against a broader range of serotypes.

Here we test the impact of this assumption by varying the two vaccine efficacy measures – efficacy against carriage and efficacy against disease. We computed the WTP threshold of PCV20 above the cost of PCV13 under the different PCV20 efficacy scenarios (Fig. 7). We show the change in the willingness to pay, relative to the default parameters for PCV20 (pink dot). In addition, to better focus on the impact of efficacy on the paediatric programme, we assumed that PPV23 vaccination in older adults continues indefinitely.

**Fig. 7.**
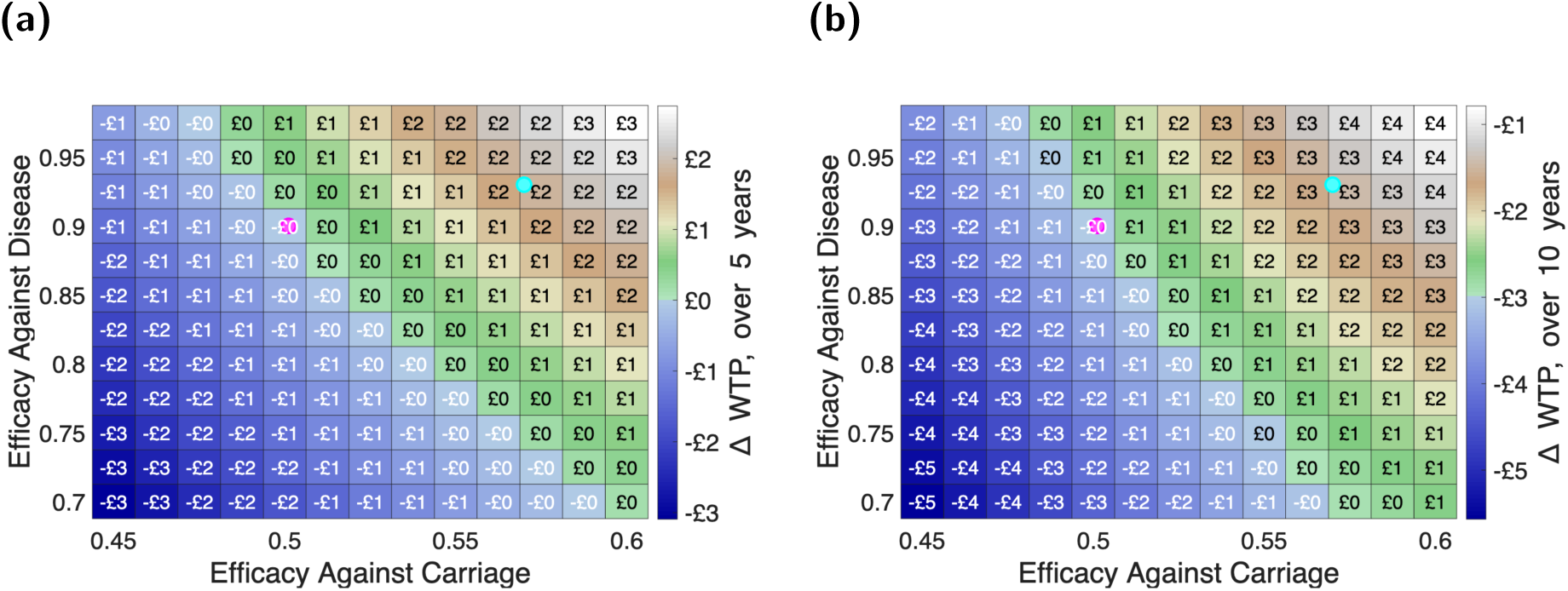
Change in the Willingness to Pay thresholds for PCV20, compared to the default PCV20 parameters (pink dot). Results are shown for two early exemplar time horizons: **(a)** 5 years; and **(b)** 10 years. Squares are colour-coded with a “geographic” colour-scheme, such that parameters associated with a lower WTP are blue and higher WTP values transition from green to brown to white. We mark with cyan and magenta dots the estimated efficacies of PCV7 and PCV13 (55% against carriage and 95% against disease for PCV7, 48% against carriage and 90% against disease for PCV13), and note that our default assuming is that PCV20 has the same efficacy as estimated for PCV13. (Throughout we continue vaccinating older adults with PPV23, to focus on the impact of PCV20 efficacy in children.)

For short time horizons, the change in the WTP threshold is relatively small even for modest changes in the two measures of efficacy. As expected, higher protection against carriage and against disease both lead to a higher threshold. Nonetheless, even for the higher efficacy parameters associated with PCV7 we only observe a £2 (at 5-years) or £3 (at 10 years) gain. At longer timescales the dynamics become far more complex, with lower efficacies leading to the re-emergence of serotypes.

## 4 Discussion

To investigate the cost-effectiveness of the pneumococcal vaccination programme in England, we have developed a parsimonious model of pneumococcal transmission and associated disease that captures the dynamics of 25 specific serotypes (plus an ‘Other’ serotype classification). The motivation for our study is the potential for the infant pneumococcal vaccination programme in England transitioning from the 13-valent PCV13 vaccine to the 20-valent PCV20 vaccine; we also consider the switch from PPV23 to PCV20 in those over 65 year old and the historic introductions of the PCV7 and PCV13 vaccines in infants. The transmission model captures three important aspects of pneumococcal epidemiology: the differential dynamics and severity of the serotypes; the competition and cross-reaction between serotypes; and the action of the vaccines on carriage and disease. The interaction of these three elements leads to serotype replacement as vaccination is introduced - driving down the cost-effectiveness of low-valent vaccines. We fundamentally believe that a serotype-specific model (with as many serotypes as practical) is needed to fully capture the complex dynamics of replacement. We match our 26-serotype model to age-structured carriage data (collected from households with young children) and disease data (invasive pneumococcal disease data for England from 2000 to 2023). We obtain a good fit to the IPD incidence over time (Fig. 3 and Fig. S5), which provides confidence in our projections.

To ascertain the cost-effectiveness of each vaccine, we utilise a health-economic approach. Specifically, we calculate willingness to pay (WTP) thresholds for each change in vaccine policy. As such, the WTP threshold for the introduction of PCV7 in 2006 is the maximal price of one vaccine dose such that the introduction of PCV7 is cost-effective (we included administration costs of £10.06 in this calculation); whereas when considering the switch to PCV13 in 2010, the WTP threshold is the maximum price of the PCV13 vaccine above the price of the PCV7 vaccine for which the switch is cost effective. These calculation relies on long-term model forecasts of the dynamics of pneumococcal infection and the associated levels of IPD (invasive pneumococcal sisease) and pCAP (pneumococcal community-acquired pneumonia). We used bespoke health economic parameter estimates for hospital costs and QALY losses for IPD, pCAP and associated sequelae to translate disease burden to an economic value. Our health economic evaluations took into account both the most likely scenario and uncertainty, ensuring these evaluations conform to JCVI guidelines (see Annex 5, Section 3 of the JCVI code of practice [23]).

Our WTP thresholds are strongly influenced by serotype replacement; as such, our estimates vary considerably with the time horizon used in the simulations. Although we are likely to see many future changes to the pneumococcal vaccination programme, we feel that longer time horizons of 25 or 50 years are the most appropriate measures. Time horizons of that scale fully capture the range of future dynamic behaviour. Our projections of IPD following the switch to PCV20 in infants showed that the reduction in IPD caused by the additional serotypes in PCV20 outweighed replacement disease from non-vaccine serotypes (Fig. 6b). These dynamics for PCV20 echo the qualitative findings by Choi *et al.* [13], who used a four serotype-group structure in their model that highlighted the role of higher case-carrier ratios for the additional serotypes covered by PCV20. Our approach takes the modelling one stage further and provides a full cost-effectiveness analysis.

We found that the WTP threshold for replacing PCV13 with PCV20 in infants is highly sensitive to the time-horizon of simulations and is also impacted by the vaccine deployed in older adults (Fig. 5). Over a five year time span, the WTP threshold for PCV20 (in addition to the price paid for PCV13) is £6 - in part due to the time required for vaccination to generate a reduction in IPD cases. This WTP estimate is comparable to the recent estimates made by Pfizer for the UK using a 5-year time horizon [14]; the authors of that study estimated PCV20 to be cost effective at £7.50 above the cost of PCV13. Over 50-year time scales (and including a 3.5% discounting) the WTP threshold increases to £84 if the elderly vaccine remains PPV23 or £76 if there is the anticipated switch to PCV20 in the elderly. This reflects the additional protection offered by PCV20 in the elderly; leading to a WTP threshold for PCV20 compared to PPV23 in the elderly of £32 (over 50 years assuming that infants are vaccinated with PCV13, or £11 if the infant programme switches to PCV20). These willingness to pay thresholds are slightly lower than recent results by Danelian *et al.* [18], who estimated a WTP threshold of £44 for PCV20 compared to PPV23 (assuming PCV13 continues in infants). Our results also reflect the efficacy of adding a second dose to the adult vaccination schedule at age 75, which is likely to become more important as we expect to see an increase in elderly populations in the future. Our estimated WTP threshold for this additional dose (excluding the extra administration fee) is very close to the list price of £56.80 for PCV20 over a 50-year time horizon.

We can also use the model retrospectively to consider the WTP thresholds for the two PCV changes that have already occurred - the introduction of PCV7 in 2006, and the switch to PCV13 in 2010-although for longer time horizons these still require the simulation of future dynamics. Compared to model projections made before the dates of introduction for each of PCV7 [11] and PCV13 [12], our formulation has the benefit of hindsight. Specifically, we are able to fit to data available after the introduction of these vaccines, meaning we can obtain insights into the importance of serotype replacement. Initial estimates of three-dose PCV7 vaccination, made in 2004, suggested it would be cost-effective at £10-15 per dose [11]. In contrast, we find that the vaccine is not cost effective at any price due to rapid serotype replacement; the willingness to pay threshold for a five-year time horizon is -£13. Finding the introduction of PCV7 vaccination in 2006 to not be cost-effective at any price is supported by a simple statistical extrapolation of the data that shows rapid replacement an a negative willingness to pay (Section S8). In 2012, van Hoek *et al.* [12] suggested that PCV13 vaccination deployed as a 3 dose (2+1 strategy) vaccine would be cost effective at £49.60 (plus a £7.50 administration fee) compared to stopping vaccination. Our results support this, predicting a willingness to pay of £60 over 50-years for a 3-dose programme compared to continuing with PCV7.

All models are abstractions of the underlying processes. There are non-trivial methodological challenges in the calibration of multi-serotype models to epidemiological data. There has to be a judicious balance in the number of individual serotypes modelled, and hence the number of parameters that need to be inferred, against the desire to fully capture historical epidemiological patterns. There is also the question of parsimony, reaching close agreement with the observed dynamics whilst fitting the minimal number of parameters. For this reason, we generally assumed that there was a simple multiplicative interaction between serotype and age (for example, the case-carrier ratio is the product of age-dependent and serotype-dependent factors), rather than attempting to fit a full matrix of values to cover all combinations of age group and serotype.

Although most of the individual serotypes in our model fit reasonably well to the temporal and age-dependent trends in IPD (Fig. 3), there were some notable discrepancies, especially in the early 2000s. These discrepancies may be partially associated with poorly understood temporal changes in the transmission dynamics that are unrelated to vaccination, such as pronounced changes in child-care, schooling, working and mixing patterns over the last 25 years.

We have assumed throughout that all serotypes are governed by a common set of equations, with only a few key parameters governing differences between serotypes. Our parsimonious approach is vital for successful parameter estimation, but may ignore fundamental epidemiological factors. For example, the data for IPD due to serotype 1 has the highest reported levels of disease in the 15-44 age group, but we are not able to capture this with our current mathematical framework as all serotypes have a common age-dependency. We note that it has been common in previous modelling studies of pneumococcal transmission applied to England to exclude serotype 1 from their analysis [13, 48].

Another potential issue is that we have amalgamated the data for multiple serotypes into an ‘Others’ group. This grouping parallels a common approach when modelling pneumococcal infection, where serotypes are amalgamated based on vaccine protection (e.g. serotypes 4, 6B, 9V, 14, 18C, 19F and 23F are modelled as a single class that is targetted by PCV7) [12, 13]. However, the amalgamating serotypes approach may camouflage the changes within this amalgamated group. It may therefore fail to anticipate the rise of one serotype that is part of this amalgamation. Specifically, a low incidence serotype that has been consigned to this ‘Other’ group could rise to prominence once PCV20 is introduced, mirroring the observed behaviour of 6C or 10A after PCV7 was introduced (whereas those serotypes had caused very little disease in the early 2000s).

There are many ways in which the modelling is severely limited by the available data. Fundamentally, the model needs to be underpinned by carriage studies that inform about infection in all ages. Although there are detailed studies of carriage in young children and their households [21, 22], there is limited information on older age groups. In addition, the only longitudinal data available (which informs about the duration of carriage) was the household study conducted in the pre-PCV7 era [21]. Moreover, for many serotypes, there are very few reported incidences of carriage [21], with serotypes 1, 4, 5, 8, 9V, 12F and 20 rarely reported despite their role in IPD (comparing Figures 3 and S4).

Similarly, data on pneumococcal community-acquired pneumonia (pCAP) is extremely limited compared to Invasive Pneumococcal Disease (IPD), with the best data associated with cases from Nottingham Hospital Trust [36, 37]. In addition, while the serotypes leading to IPD are well recorded, there are few studies that link severity and sequelae to serotypes [49, 50]. Although these studies provide snap-shots of the dynamics in particular locations, there remain questions about extrapolation to the national scale. We have therefore chosen not to include additional measures of serotype severity in our model.

Finally, there are limitations imposed by the sparsity of data on vaccines and levels of real-world protection. Assumptions about the protective impacts of PCV20 are largely based on immunological comparisons with PCV7 and PCV13. Information on the duration of protection from all vaccines is also extremely limited and difficult to collect. It is important to have ongoing monitoring of real-world vaccine effectiveness against targeted seroptypes, with estimates per serotype where possible.

This work has demonstrated the capability of pneumococcal infection models that capture the interaction between multiple serotypes for understanding serotype replacement. With hindsight and the benefit of additional years of data, we conclude that the introduction of PCV7 vaccination for infants in 2006 in England was unlikely to be cost-effective. On the other hand, the prospective switch from PCV13 to PCV20 is cost-effective providing PCV20 is not too expensive (costing less than £76 more than PCV13). Similarly, we find that the replacement of PPV23 vaccination in those over 65 year of age with a single dose of PCV20 is also cost effective, although at a lower willingness to pay threshold. Given England has an ageing population, we believe that a booster dose of PCV20 at age 75 may also be cost-effective (depending on the vaccine price) due to the waning of vaccine protection and the burden of disease in the very elderly. We stress, however, that these projections should be treated with caution; it has not been possible to model all known serotypes (both computationally and due to data availability). It is feasible that novel (non-modelled) serotypes could arise, dominate, and invalidate our predicted dynamics.

## Author contributions

**Matt J. Keeling:** Conceptualisation, Data curation, Formal analysis, Funding acquisition, Methodology, Software, Supervision, Validation, Visualisation, Writing - Original Draft, Writing - Review & Editing.

**Omar El Deeb:** Data curation, Formal analysis, Methodology, Software, Validation, Visualisation, Writing - Original Draft, Writing - Review & Editing.

**Phuong Bich Tran:** Data curation, Formal analysis, Methodology, Writing - Review & Editing.

**Stavros Petrou:** Funding acquisition, Methodology, Writing - Review & Editing.

**Edward M. Hill:** Conceptualisation, Data curation, Formal analysis, Funding acquisition, Methodology, Software, Supervision, Visualisation, Writing - Original Draft, Writing - Review & Editing.

## Data Availability

All data produced in the present study are available upon reasonable request to the authors

## Acknowledgements

The authors express their gratitude to Professor Wei Shen Lim of Nottingham University Hospitals NHS Trust and Nottingham Biomedical Research Centre, and Dr. Louise Lansbury of University of Nottingham and Nottingham Biomedical Research Centre for providing additional data from the Study of Community-Acquired Pneumonia (SCAPA). We are also extremely grateful to Prof Elizabeth Miller (UKHSA) for providing access to the five UK carriage studies undertaken from 2021 to 2018. We thank Prof Shamez Ladhani and Dr Fariyo Abdullahi (UKHSA) for their assistance in providing the data on IPD cases in England. Finally, we thank the JCVI pneumococcal subcommittee, especially Prof Caroline Trotter (Cambridge) and Stefan Flasche (LSHTM), for their helpful thoughts and comments during the development of this work.

