## Supplementary Information for "A model-based evaluation of the cost-effectiveness of paediatric and elderly vaccination against pneumococcal infection in England"

**S5 Model fitting to IPD data** **15**

**S6 Model fitting to CAP data** **16**

**S7 Health economic model parameterisation** **18**

**S8 Statistical fits to IPD data** **23**

### S1 Data Sources

#### S1.1 Demographics

We used three sources of demographic data to capture population changes in England over time: single year of age population estimates for England from the 2021 census, provided by the Office for National Statistics [1]; age-group estimates for the period 2000-2020 formulated by ONS and based on census data; and Office for National Statistics projected population estimates for England for the respective future years [2].

#### S1.2 Epidemiological outcomes

**Carriage:** We used two sets of age structured and serotype dependent carriage data. The first was obtained from a longitudinal household carriage study conducted before the introduction of PCV7 [3]. The carriage study sampled families that had young children in the year of 2001/2002. Each individual in the family was tested for carriage for 10 consecutive months. The monthly test results (together with the age of the individual and the individual/household identifier) were reported as either negative, positive with a specific serotype (or serotypes) or positive with an unspecified serotype. The second sets of carriage data were based on four subsequent studies undertaken in 2008, 2012, 2015 and 2018, using a similar sample of households with young children, but without repeated sampling of the same individual [4]. The sample dates for these data span a time period after the introduction of PCV7 (the 2008 sampling) to after the introduction of PCV13 (the 2012, 2015 and 2018 samplings). In total data were available on approximately 2400 individuals, although there is a strong bias towards younger age-groups.

**IPD** The UK Health Security Agency (UKHSA) (and before that Public Health England (PHE)) monitors IPD cases in hospitals. In each calender year, cases are stratified into six age-groups (under 2, 2-4, 5-14, 15-44, 45-64 and over 65 years old), serotype (where available, with over 90% having their serotype determined since 2010) and the total number of IPD cases. As such the IPD data contains over 114,000 serotyped cases between 2000 and 2023.

**pCAP** Relative to data on IPD, data on pneumococcal CAP is limited. Many studies have focused on CAP in general without specifying its particular aetiology. A major study was conducted on adults over 16 years old hospitalised with CAP between 2013 and 2023 in two hospitals in Nottingham [5]. It covered 5186 patients with CAP among which 2193 had pneumococcal pneumonia and analysed their pneumococcal serotypes. The final form of the available data is serotype- and year-specific, but not stratified by age. We inferred the age distribution of pCAP from two studies. For adults, we utilised a sample of 920 adult patients with CAP in the Nottingham region over the years of 2008-2010, among which 366 patients were identified with pCAP [6]. The risk of pCAP in children aged 15 or under is based on a reported incidence rate of CAP estimated to be 144 in $100,000$ [7], of which 46% (66 per $100,000$ are due to pneumococcal infection [8]. We further assumed the age-distribution of pCAP in children to have the same profile as IPD.

#### S1.3 Vaccination uptake

To inform the uptake of pneumococcal vaccines in childhood age groups, we used childhood immunisation programme data reported by the Cover of Vaccination Evaluated Rapidly (COVER) and the NHS Immunisation Statistics resources for the time period 2006 to 2023 [9, 10]. COVER annual reports contain the percentage of the primary course of PCV received by the first birthday and percentage of the secondary course of PCV (i.e. the booster dose) received by the second birthday. The annual reporting year runs from 01 April to 31 March in the subsequent calendar year. The NHS annual reports contain the percentages of primary courses completed with the vaccine by the first birthday. We adopted a rounded figure of 93% vaccine uptake from 2006 to 2010, followed by a 91% uptake from 2010 onwards. For all projections, we assumed a vaccine uptake of 91%.

The annual reports of the Department of Health and Social Care (DHSC) [11] show that the PPV23 vaccine coverage amongst adults 65 years or older initially started at 65% in 2005, but has consistently maintained a coverage of 69% to 71% between 2006 and 2021. More recent data suggests a slight rise to 73% in 2023/2024 [12]. From these data, we assumed a flat average uptake of PPV23 of 72%. We assumed the same uptake of 72% for scenarios where PPV23 was replaced by PCV20 in the adult pneumococcal vaccination programme.

##### S1.4 Vaccination efficacy

The pneumococcal vaccines, including both PCV7 and PCV13, are more effective against disease (both IPD and pCAP) than against carriage. Data from England and Wales suggest that although there are some variation with serotype, overall vaccine efficacy against disease for PCV7 ($VE_D^{\mathrm{PCV7}}$) was about 93% for fully vaccinated children Andrews *et al.* [13] PCV13 targets a broader set of serotypes, and so effectiveness against each serotype is reduced. Aside from serotype 3, which shows vaccine escape, the PCV13 efficacy against IPD was around 90% [14] for fully vaccinated children at $< 2$ years of age, although again there is some serotype variability.

A meta-analysis of ten studies found that efficacy against carriage was estimated to be 57% for PCV7 at 6 months after completion of the vaccination schedule [15], and this is the point estimate we use throughout this work. For PCV13 there exists only two studies, which estimate efficacy against carriage between 62-65% for fully vaccinated individuals at 24 months of age [16, 17]. However, the model inference using English data suggested that the efficacy of PCV13 was far lower Choi *et al.* [18]. For our baseline assumption we assumed PCV20 to have similar characteristics to PCV13. We also assume that PCV20 in older adults has the same efficacy against carriage and disease as it does in infants. We treated the duration of vaccine-derived protection as a free parameter that we estimated as part of our fitting procedure.

PPV23 has been found to be effective against IPD, but ineffective against carriage and pCAP [19]. Our estimates of PPV23 efficacy against IPD are extrapolated from observations of 41% for those vaccinated within two years, to 34% for those vaccinated 2–4 years earlier, and 23% for those vaccinated 5 or more years ago Djennad *et al.* [20].

#### S2 Pneumococcal transmission model description

There being many pneumococcal serotypes poses challenges to traditional compartmental-type infectious disease models in producing a tractable individual-serotype transmission model for pneumococcal carriage and disease.

We instead adopted a different model structure, based on the multi-strain pathogen model formulation presented by Gog and Grenfell [21]. In this model framework, a deterministic system of ordinary differential equations tracks for each serotype how many people are susceptible (see Section S2.1), vaccinated (see Section S2.2), infected (see Section S2.3) or resistant (see Section S2.4) to that serotype.

We considered the pneumococcal serotypes:

- Contained in PCV7 vaccine: 4, 6B, 9V, 14, 18C, 19F, 23F
- Additional serotypes in PCV13 vaccine: 1, 3, 5, 6A, 7F, 19A

- Additional serotypes in PCV20 vaccine: 8, 10A, 11A, 12F, 15B, 22F, 33F
- Not contained in PCV7/PCV13/PCV20 vaccines: 2, 6C, 9N, 17F, 20, Others

As well as stratifying the population by serotype, we also stratified the population into nine age groups: 0 up to 3 months, 3 up to 12 months, 12 months up to 2 years, 2-4 years, 5-14 years, 15-44 years, 45-64 years, 65-75 years, 75+ years. From the transmission model we generated multiple age- and serotype-specific epidemiological measures (more details in Section S2.5).

#### S2.1 Susceptible states and force of infection

The number of people aged $a$ and susceptible to serotype $i$ obeys:

$$\begin{aligned} \frac{dS_{a,i}}{dt} &= B_a - \lambda_{a,i} S_{a,i} \left[1 - \sum_{j \neq i} I_{a,j}/N_a\right] - \lambda_{a,i} S_{a,i} \left[\sum_{j \neq i} C_{i,j} I_{a,j}/N_a\right] \\ &+\omega_i m R^m_{a,i} + \Omega_i m \sum_{VT} V^{VT,m}_{a,i} - \mu_a S_{a,i} + \mu_{a-1}(1 - \widehat{v}_{a,i}) S_{a-1,i} \\ &- \sum_{k \neq i} Z_{i,k} \gamma_{a,k} I_{a,k} S_{a,i}/(N_a) \end{aligned}$$

The first term is the birth rate $B_a$ and is only non-zero for the first age-group. The second term is infection of susceptible individuals that are not infected with another strain; the third term is infection of susceptibles who are currently infected with another strain; the fourth and fifth terms are the waning of immunity obtained from natural infection and vaccination, respectively. The sixth and the seventh terms capture the demographic impact of continuous aging (and vaccination that occurs at the boundary of two age groups). The eighth term is immunity to serotype $i$ being acquired via recovery to serotype $k$ that induces cross-immunity to serotype $i$.

For most serotypes $Z_{i,j}$ is zero. There are, however, the following exceptions: the interaction between type 6 serotypes (6A, 6B and 6C); the interaction between 8, 9N, 12F and 22F; and the interaction between 19A and 19F. We inferred the strength of $Z$ for these serotype combinations. The competition matrix $C_{i,j}$ determines how much the presence of serotype $j$ affects the infection with serotype $i$; when $C_{i,j} = 1$ serotype $i$ is unaffected by serotype $j$. Rather than attempting to infer the entire $26 \times 26$, we define a fitness $F_i$ for each serotype. We use this fitness vector to generate our matrix

$$C_{i,j} = \begin{cases} \max\left(1 - \frac{F_j}{F_i}, 0\right) & \text{if } Z_{i,j} = 0 \\ 0 & \text{otherwise.} \end{cases}$$

This ensures that whichever serotype has maximal fitness can invade an already infected individual, with the exception being when serotypes strongly interact.

We define the age-based force of infection, $\lambda_{a,i}$, as:

$$\lambda_{a,i} = \beta_{a,i} \sum_b M_{a,b} I_{b,i}/N_b$$

where $M$ is the who acquires infection from whom mixing matrix.

##### S2.2 Vaccination states

The equations governing the rate of change in the vaccinated state population sizes had a similar structure to the susceptible state rate of change equations. For completeness, we also separate the vaccine compartments by the type of vaccine received ($VT = \{$PCV7, PCV13, PCV20 or PPV23$\}$):

$$\begin{aligned}
\frac{dV_{a,i}^{VT,1}}{dt} &= \mu_{a-1}v_{a,i}^{VT}S_{a-1,i} - \lambda_{a,i}\rho_i^{VT}V_{a,i}^{VT,1}\left[1-\sum_{j\neq i}I_{a,j}/N_a\right] - \lambda_{a,i}\rho_i^{VT}V_{a,i}^{VT,1}\left[\sum_{j\neq i}C_{i,j}I_{a,j}/N_a\right] \\
&\quad -\Omega_i^{VT}mV_{a,i}^{VT,1} - \mu_a V_{a,i}^{VT,1} + \mu_{a-1}V_{a-1,i}^{VT,1} - \sum_{k\neq i}Z_{i,k}\gamma_{a,k}I_{a,k}V_{a,i}^{VT,1}/(N_a) \\
\frac{dV_{a,i}^{VT,n}}{dt} &= \Omega_i^{VT}mV_{a,i}^{VT,n-1} - \lambda_{a,i}\rho_i^{VT}V_{a,i}^{VT,n}\left[1-\sum_{j\neq i}I_{a,j}/N_a\right] - \lambda_{a,i}\rho_i^{VT}V_{a,i}^{VT,n}\left[\sum_{j\neq i}C_{i,j}I_{a,j}/N_a\right] \\
&\quad -\Omega_i^{VT}mV_{a,i}^{VT,n} - \mu_a V_{a,i}^{VT,n} + \mu_{a-1}V_{a-1,i}^{VT,n} - \sum_{k\neq i}Z_{i,k}\gamma_{a,k}I_{a,k}V_{a,i}^{VT,n}/(N_a)
\end{aligned}$$

with a parameter $\rho$ capturing if the vaccine leads to complete or partial immunity (vaccine efficacy against carriage $= VE_C^X = 1-\rho^X$), and the integer parameter $m$ enabling flexibility in the model based on whether the waning of vaccine induced immunity is best captured as Markovian (where $m = 1$) or is best captured by an Erlang distribution (with $m$ an integer greater than 1).

To simplify the following equations we define the total susceptibility of the vaccinated population to be:

$$\widehat{V}_{a,i} = \sum_{VT,n}\rho_i^{VT}V_{a,i}^{VT,n}$$

and the total rate of vaccination is:

$$\widehat{v}_{a,i} = \sum_{VT}v_{a,i}^{VT}.$$

where $\rho_i^{VT} = \rho^{VT} \times \rho_i$.

##### S2.3 Infection (carriage) states

We model infection (carriage) as:

$$\begin{aligned}
\frac{dI_{a,i}}{dt} &= \lambda_{a,i}S_{a,i}\left[1-\sum_{j\neq i}I_{a,j}/N_a\right] + \lambda_{a,i}S_{a,i}\left[\sum_{j\neq i}C_{i,j}I_{a,j}/N_a\right] \\
&\quad +\lambda_{a,i}\widehat{V}_{a,i}\left[1-\sum_{j\neq i}I_{a,j}/N_a\right] + \lambda_{a,i}\widehat{V}_{a,i}\left[\sum_{j\neq i}C_{i,j}I_{a,j}/N_a\right] \\
&\quad -\sum_{j\neq i}\lambda_{a,j}I_{a,i}C_{j,i}\left[S_{a,j}+\widehat{V}_{a,j}\right]/N_a - \gamma_{a,i}I_{a,i} \\
&\quad -\mu_a I_{a,i} + \mu_{a-1}I_{a-1,i}
\end{aligned}$$

Here we have assumed there is just a single infectious class - leading to exponential duration of infection. The first two terms are the same as in the susceptible model; the second are from vaccinated (and partially immune) class. The fifth term is about people infected with serotype $i$ getting infected with serotype $j$, which dominates and out competes $i$. The sixth term is recovery, which we assumed to occur at a serotype dependent rate (it could also depend on age). The final two terms again relate to continuous ageing. (When dealing with disease states - IPD and pCAP - we further partition the infected population by whether the individual was susceptible or in a particular vaccination class at the time of infection).

##### S2.4 Recovered states

Finally, we assumed that immunity wanes over time.

$$\begin{aligned}
\frac{dR^1_{a,i}}{dt} &= \gamma_{a,i}I_{a,i} + \sum_{j\neq i}\lambda_{a,j}I_{a,i}C_{j,i}\left[S_{a,j} + \widehat{V}_{a,j}\right]/N_a + \sum_{k\neq i} Z_{i,k}\gamma_{a,k}I_{a,k}(N_a - R^1_{a,i} - I_{a,i})/N_a \\
&\quad -\omega_i m R^1_{a,i} - \mu_a R^1_{a,i} + \mu_{a-1}R^1_{a-1,i} \\
\frac{dR^n_{a,i}}{dt} &= \omega_i m R^{n-1}_{a,i} - \omega_i m R^n_{a,i} - \sum_{k\neq i} Z_{i,k}\gamma_{a,k}I_{a,k}R^n_{a,i}/(N_a) - \mu_a R^n_{a,i} + \mu_{a-1}R^n_{a-1,i}
\end{aligned}$$

where $n = 1, \ldots, m$ and $Z_{i,k}$ captures immunity to serotype $i$ due to infection with serotype $k$. Here the first equation has terms for the natural loss of susceptibility and for the replacement of a serotype by a more dominant serotype. Note that in this formulation we assumed that immunity from infection ($R$) was stronger than immunity from vaccination ($V$); we therefore did not associate the vaccination of recovered individuals with any transition in states.

##### S2.5 Model outputs

From the pneumococcal transmission model, we output two epidemiological measures through time: (i) carriage of each serotype for those in age-group $a$ which is $I_{a,i}$; and (ii) IPD (invasive pneumococcal disease), calculated as $D_{a,i}I_{a,i}$ where the risk of disease (or case-to-carrier ratio) $D_{a,i}$ is dependent on both age and serotype.

#### S3 Model Cross Comparison

The model used throughout this work is a low-dimensional simplification of the full compartmental model in which every possible state is explicitly modelled. To test the agreement of these two approaches, we consider a model with just two pneumococcal serotypes ($i = 1, 2$), and assume that the vaccine only protects against serotype 1 ($\rho_1 < 1$) with serotype 2 unaffected ($\rho_2 = 1$).

##### S3.1 Structured model

Using the same notation as for the 26-serotype model we have:

$$\begin{aligned}
\frac{dS_1}{dt} &= B(1-v_1) - \lambda_1 S_1\left(1-\frac{I_2}{N}\right) - \lambda_1 S_1 C_{1,2}\frac{I_2}{N} + \omega_1 R_1 + \Omega_1 V_1 - \mu S_1 - Z_{1,2}\gamma_2 I_2 \frac{S_1}{N}\\
\frac{dS_2}{dt} &= B - \lambda_2 S_2\left(1-\frac{I_1}{N}\right) - \lambda_2 S_2 C_{2,1}\frac{I_1}{N} + \omega_2 R_2 - \mu S_2 - Z_{2,1}\gamma_1 I_1 \frac{S_2}{N}\\
\frac{dV_1}{dt} &= v_1 B - \lambda_1\rho_1 V_1\left(1-\frac{I_2}{N}\right) - \lambda_1\rho_1 V_1 C_{1,2}\frac{I_2}{N} - \Omega_1 V_1 - \mu V_1 - Z_{1,2}\gamma_2 I_2\frac{V_1}{N}\\
\frac{dV_2}{dt} &= 0\\
\frac{dI_1}{dt} &= \lambda_1 S_1\left(1-\frac{I_2}{N}\right) + \lambda_1 S_1 C_{1,2}\frac{I_2}{N} + \lambda_1\rho_1 V_1\left(1-\frac{I_2}{N}\right) + \lambda_1\rho_1 V_1 C_{1,2}\frac{I_2}{N}\\
&\quad -\lambda_2 I_1 C_{2,1}\frac{S_2+\rho_2 V_2}{N} - \gamma_1 I_1 - \mu I_1\\
\frac{dI_2}{dt} &= \lambda_2 S_2\left(1-\frac{I_1}{N}\right) + \lambda_2 S_2 C_{2,1}\frac{I_1}{N} - \lambda_1 I_2 C_{1,2}\frac{S_1+\rho_1 V_1}{N} - \gamma_2 I_2 - \mu I_2\\
\frac{dR_1}{dt} &= \gamma_1 I_1 + \lambda_2 I_1 C_{2,1}\frac{S_2+\rho_2 V_2}{N} - \omega_1 R_1 - \mu R_1 + Z_{1,2}\gamma_2 I_2 (N - R_1 - I_1)/N\\
\frac{dR_2}{dt} &= \gamma_2 I_2 + \lambda_1 I_2 C_{1,2}\frac{S_1+\rho_1 V_1}{N} - \omega_2 R_2 - \mu R_2 + Z_{2,1}\gamma_1 I_1 (N - R_2 - I_2)/N
\end{aligned} \tag{1}$$

###### S3.1.1 Analytical Solutions and stability criteria

Given the relative simplicity of the equations with just two serotypes, we can seek an analytical understanding of the dynamics and competition. In particular, we can perform the following calculations: finding the equilibrium of serotype 1 alone with and without vaccination; finding whether serotype 2 can invade when serotype 1 is as equilibrium. Using these calculations we can then find regions of parameter space in which serotype 1 dominates in the absence of vaccination, but serotype 2 invades and serotype 1 is driven to extinction once vaccination is applied.

Without vaccination, our non-trivial endemic equilibrium is:

$$S_1^* = \frac{N(\gamma_1+\mu)}{\beta_1} = \frac{N}{R_{0,1}}, \quad I_1^* = \frac{\big(B\beta_1 - N\mu(\gamma_1+\mu)\big)(\mu+\omega_1)}{\beta_1\mu(\gamma_1+\mu+\omega_1)}, \quad R_1^* = \frac{B\beta_1\gamma_1 - N\gamma_1\mu(\gamma_1+\mu)}{\beta_1\mu(\gamma_1+\mu+\omega_1)}.$$

which is stable as long as the basic reproductive ratio associated with serotype 1 is greater than 1:

$$R_{0,1} = \frac{\beta_1}{\gamma_1+\nu} > 1 \tag{2}$$

Setting $W_1 = \omega_1 + \mu$, the equilibrium can be simplified to

$$S_1^* = \frac{N}{R_{0,1}}, \quad I_1^* = N\left(1-\frac{1}{R_{0,1}}\right)\frac{W_1}{W_1+\gamma_1}, \quad R_1^* = N\left(1-\frac{1}{R_{0,1}}\right)\frac{\gamma_1}{W_1+\gamma_1}.$$

When vaccination is introduced into this single serotype model the equilibrium state becomes much more complex:

$$S_1^* = N\left(\frac{1}{R_{0,1}} - \frac{\mu\,\rho_1 v_1}{\rho_1\lambda_1^* + \Theta_1}\right), \qquad V_1^* = N\frac{\mu\, v_1}{\rho_1\lambda_1^* + \Theta_1}$$

where $\Theta_1 = \Omega_1 + \mu$. While the infectious and recovered can be written in terms of the equilibrium force of infection ($\lambda_1^*$):

$$I_1^* = \frac{\lambda_1^* N}{\beta_1}, \quad R_1^* = \frac{\gamma_1}{W_1} I_1^*.$$

The equilibrium force of infection is the larger (positive root) solution to the quadratic equation:

$$\left[\rho_1 \frac{W_1+\gamma_1}{W_1}\right]\lambda^2 + \left[\frac{W_1+\gamma_1}{W_1}\Theta_1 - \rho_1(\beta_1 - A_1)\right]\lambda + \left[-\Theta_1(\beta_1 - A_1) + \beta_1\,\mu\, v_1(1-\rho_1)\right] = 0$$

While these analytic expressions are too complex to provide much intuitive insight, we find that the solution is only non-zero, real and stable if:

$$R_{0,1}\left[1 - \frac{(1-\rho_1)\,\mu\, v_1}{\Theta_1}\right] > 1$$

When this term is less than one it provides a condition for vaccination to eradicate serotype 1:

$$v_1 > \Theta_1 \frac{1 - \frac{1}{R_{0,1}}}{\mu(1-\rho_1)} \tag{3}$$

Considering the invasion of serotype 2, when serotype 1 is at equilibrium, we show that for small $I_2$:

$$\frac{dI_2}{dt} = \left[\beta_2 \frac{S_2^*}{N}\left(1 - (1 - C_{21})\frac{I_1^*}{N}\right) - \left(\gamma_2 + \mu + C_{12}(\gamma_1 + \mu)\frac{I_1^*}{N}\right)\right] I_2.$$

noting that even when serotype 2 is not present, $S_2^*$ can be reduced by the cross-protection between serotypes as captured by $Z_{21}$:

$$S_2^* = \frac{(\omega_2 + \mu)N^2}{(\omega_2 + \mu)N + Z_{21}\gamma_1 I^*}$$

The invasion depends on the relative size of the two terms within the growth rate expression, allowing us to define an invasion threshold, which must be greater than 1 for invasion:

$$\mathcal{R}_2^{\text{inv}}(I_1^*) := \frac{\beta_2 S_2^*\left[N - (1 - C_{21})I_1^*\right]}{(\gamma_2+\mu)N^2 + C_{12}(\gamma_1+\mu)N I_1^*} = R_{0,2}\frac{S_2^*}{N}\frac{N - (1-C_{21})I_1^*}{N + C_{12}\frac{\gamma_1+\mu}{\gamma_2+\mu} I_1^*} > 1 \tag{4}$$

If we are interested in replacement dynamics, where before vaccination serotype 1 dominated (and serotype 2 could not invade), while after vaccination serotype 1 was driven extinct allowing serotype 2 to invade, then (based on Eq. 2-4) this is represented by the four conditions:

$$\begin{array}{rclrcl} R_{0,1} & > & 1 & \mathcal{R}_2^{\text{inv}}(I_1^*) & < & 1 \\ R_{0,1}\left[1 - \frac{(1-\rho_1)\,\mu\, v_1}{\Theta_1}\right] & < & 1 & R_{0,2} & > & 1 \end{array} \tag{5}$$

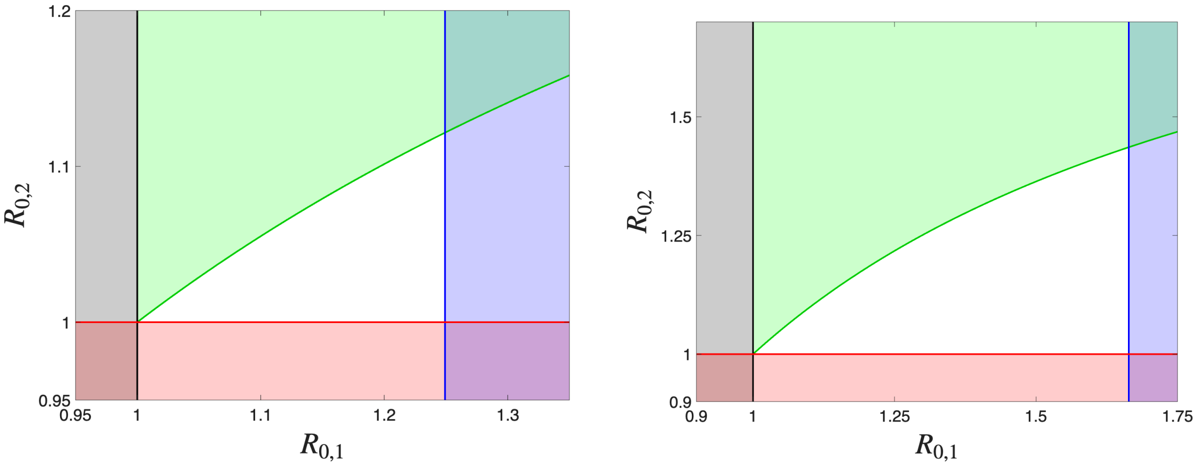

**Fig. S1.** $R_0$ parameter space plot with shaded regions showing the $(R_{0,1}, R_{0,2})$ values that violate the inequalities in Eq. (5). The vertical lines (black and blue) correspond to inequalities in Eq. (2) and Eq. (3), while the horziontal red line corresponds to $R_{0,2} > 1$ and the green curve corresponds to Eq. (4). Parameter sets are given in Table S1, with parameter set 1 on the left and 2 on the right.

These are shown in Fig. S1, with the four conditions leading to a triangular area of parameter space (white) where we expect to observe vaccine driven replacement.

We choose two sets of different parameters that conform with the parameter ranges estimated for the full model of 26-serotypes. For parameter set 1, we consider a scenario with strong competition during infection ($C_{12} = 1$) but weak cross immunity ($Z_{12} = Z_{21} = 0.3$); for parameter set 2 we focus only on the younger age-group and hence have a higher demographic turn-over, we also include weaker competition but stronger cross immunity.

| Parameter | $\mu$ | $v_1$ | $\rho_1$ | $C_{12}$ | $C_{21}$ | $Z_{1,2}$ | $Z_{2,1}$ | $\gamma_1$ | $\gamma_2$ | $\omega_1$ | $\omega_2$ | $\Omega_1$ |
|---|---|---|---|---|---|---|---|---|---|---|---|---|
| Set 1 | 1/60 | 0.95 | 0.16 | 1 | 0 | 0.3 | 0.3 | 10 | 3 | 0.09 | 0.04 | 0.05 |
| Set 2 | 1/10 | 0.95 | 0.16 | 0.5 | 0 | 0.9 | 0.9 | 10 | 20 | 0.08 | 0.05 | 0.1 |

**Table S1.** Summary of parameters used in this section. Giving values of the birth rate ($\mu$), vaccination level ($v$), vaccine protection ($\rho$), replacement coefficients ($C$), cross immunity parameters ($Z$), recovery rates ($\gamma$), natural immunity waning rate ($\omega$) and waning rate following vaccination ($\Omega$).

#### S3.2 Compartmental model

In this model formulation, we return to the traditional compartmental approach where individuals are explicitly and uniquely categorised by their status with respect to both serotype 1 and serotype 2 (e.g., $N_{SI}$ represents individuals susceptible to serotype 1 and infected with serotype 2). This formulation facilitates the precise modeling of co-infection dynamics, serotype competition, and cross immunity, each of which involves a transition in the status of both serotypes. This compartmental model allows us to test our lower-dimensional structural model.

$$
\begin{aligned}
\frac{dN_{SS}}{dt} &= B(1-v_1) - (\lambda_1+\lambda_2+\mu)N_{SS} + \omega_1 N_{RS} + \omega_2 N_{SR} + \Omega_1 N_{VS} \\
\frac{dN_{IS}}{dt} &= (1-Z_{2,1})\lambda_1 N_{SS} + \rho_1\lambda_1 N_{VS} - (C_{2,1}\lambda_2+\gamma_1+\mu)N_{IS} + \omega_2 N_{IR} \\
\frac{dN_{RS}}{dt} &= \gamma_1 N_{IS} - (\lambda_2+\omega_1+\mu)N_{RS} + \omega_2 N_{RR} \\
\frac{dN_{VS}}{dt} &= B\,v_1 - (\rho_1\lambda_1+\lambda_2+\Omega_1+\mu)N_{VS} + \omega_2 N_{VR} \\
\frac{dN_{SI}}{dt} &= (1-Z_{1,2})\lambda_2 N_{SS} + \omega_1 N_{RI} + \Omega_1 N_{VI} - (C_{1,2}\lambda_1+\gamma_2+\mu)N_{SI} \\
\frac{dN_{II}}{dt} &= -(\gamma_1+\gamma_2+\mu)N_{II} \qquad (6) \\
\frac{dN_{RI}}{dt} &= C_{2,1}\lambda_2 N_{IS} + Z_{1,2}\lambda_2 N_{SS} + \gamma_1 N_{II} + \lambda_2 N_{RS} - (\omega_1+\gamma_2+\mu)N_{RI} \\
\frac{dN_{VI}}{dt} &= \lambda_2 N_{VS} - (C_{1,2}\rho_1\lambda_1 + Z_{1,2}\gamma_2 + \gamma_2 + \Omega_1 + \mu)N_{VI} \\
\frac{dN_{SR}}{dt} &= \gamma_2 N_{SI} + \omega_1 N_{RR} + \Omega_1 N_{VR} - (\lambda_1+\omega_2+\mu)N_{SR} \\
\frac{dN_{IR}}{dt} &= C_{1,2}\lambda_1 N_{SI} + Z_{2,1}\lambda_1 N_{SS} + C_{1,2}\rho_1\lambda_1 N_{VI} + \lambda_1 N_{SR} + \rho_1\lambda_1 N_{VR} + \gamma_2 N_{II} - (\gamma_1+\omega_2+\mu)N_{IR} \\
\frac{dN_{RR}}{dt} &= \gamma_1 N_{IR} + \gamma_2 N_{RI} - (\omega_1+\omega_2+\mu)N_{RR} \\
\frac{dN_{VR}}{dt} &= \gamma_2 N_{VI} - (\rho_1\lambda_1+\Omega_1+\omega_2+\mu)N_{VR}
\end{aligned}
$$

In the state-structured model, the force of infection for each serotype is computed by summing over all compartments where the individual is currently infectious with that serotype. Assuming homogeneous mixing and no age or contact structure, we define:

$$
\begin{aligned}
\lambda_1 &= \beta_1 \cdot \frac{N_{IS}+N_{II}+N_{IR}}{N} \\
\lambda_2 &= \beta_2 \cdot \frac{N_{SI}+N_{II}+N_{RI}+N_{VI}}{N}
\end{aligned}
$$

We are now in a position to compare the results of the two model formations, albeit for a reduced number of serotypes. Fig. S2 shows simulations from the two parameter sets, with values of $R_0$ chosen such that we observe serotype replacement upon vaccination. For parameter set 1, we observe the somewhat counter intuitive phenomena that the level of carriage after replacement is comparable with the level before. In both examples we observe large long-term oscillations, which are partially attributable to invasion of the second serotype from very low levels. Although the agreement between the two models is excellent, there is a slight temporal shift in the second example which is attributable to the time taken for infection to establish from very low levels at invasion.

We can also compare these ODE simulations with the results from our theoretical stability calculations (Eq. (5)). Fig. S3, shows the four boundaries of the inequalities (as in Fig. S1), with serotype replacement calculated directly from the ODEs (showing where serotype 1 is excluded by vaccination and where serotype 2 is unable to invade when serotype 1 is at equilibrium). Magenta dots show where

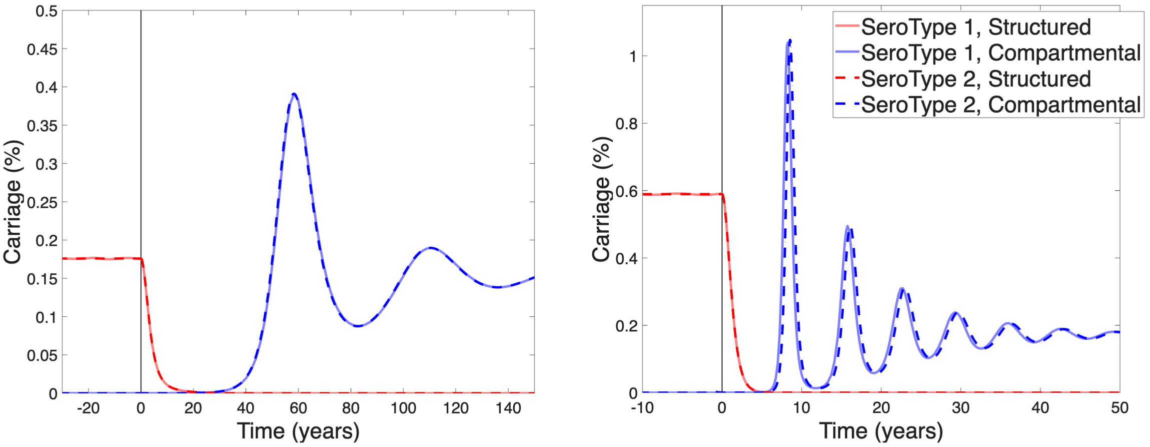

**Fig. S2.** Carriage of serotype 1 (red) and 2 (blue), before and after vaccination at time zero. Results from the structured model (pale lines) and the compartmental model (darker dashed lines ) are superimposed so that differences are clear. Left figure is for parameter set 1, right figure is for parameter set 2.

sampled parameters in $R_0$-space demonstrated serotype replacement for the structured model (Section S3.1); unsurprisingly this agrees with our analytic boundaries. Open circles show where serotype replacement occurs for the compartmental model (Eq. (6)). For the second parameter set, where the amount of cross reaction ($Z$) is relatively large, we see some disagreements in which regions of $R_0$-space are associated with serotype replacement; in particular larger values of $R_{0,2}$ still lead to exclusion of the serotype 2 due to the strong cross-correlation between protection against both serotypes.

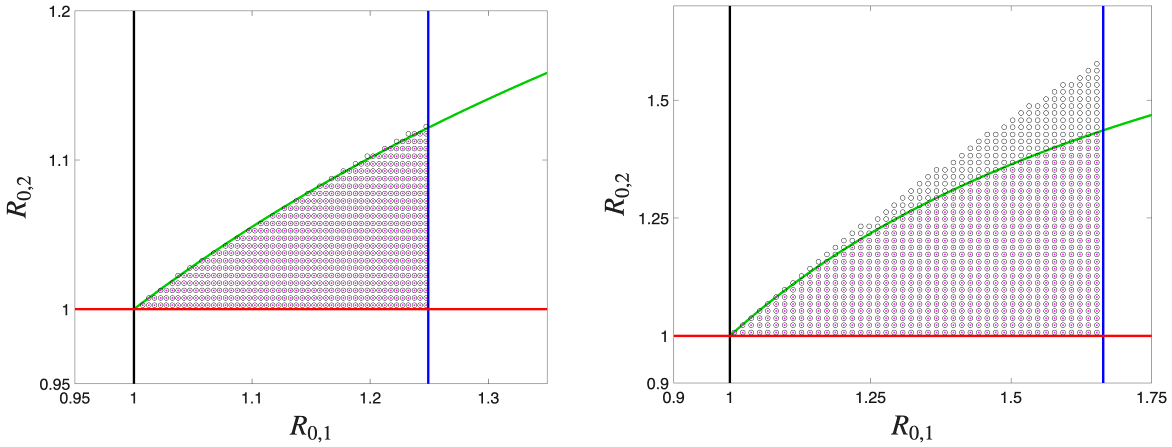

**Fig. S3.** $R_0$ parameter space plot with boundaries of the four inequalities as in Fig. S1. Magenta dots are points in parameter space where the structured ODEs display vaccine driven replacement; black circles are where the compartmental ODEs display vaccine driven replacement.

### S4 Model fitting to carriage data

We had carriage data from two sets of age structured and serotype dependent studies.

One data set was a longitudinal household carriage study conducted before the introduction of PCV7 [3]. The carriage study sampled families that had children in the year of 2001/2002. The second set of carriage data was based on cross-sectional samples taken in the years of 2008, 2012, 2015 and 2018 [4].

The data format was dependent on the time period. Per time period, we consequently constructed differing formulations for the likelihood of the model outputs compared to the observed data.

For sampling periods in 2001/2002, there was longitudinal sampling (fitting procedure to these data described in Section S4.1). For sampling periods after 2002, the data used were from cross-sectional studies, with no repeated sampling of the same individuals (fitting procedure to these data described in Section S4.2).

#### S4.1 Sampling periods 2001/2002

For this time period, we used data from a longitudinal household carriage study that sampled families with children [3]. Each individual in the family of age $a$ was tested for carriage over 10 consecutive months. The result was reported as either negative, positive with a specific serotype or positive with an unspecified serotype.

Suppose the sampled individual (age $a$) was free of serotype $i$ at time $t_1$, was first found to carry serotype $i$ at time $t_2$, was last found to carry the serotype at time $t_3$, and was clear of the serotype at time $t_4$. This implies the individual acquired infection between times $t_1$ and $t_2$; hence

$$Pr_{a,i}^{\text{inf}} = 1 - \exp\left(-\int_{t_1}^{t_2} \frac{T_{a,i}(t)}{N_a} dt\right),$$

where $T_{a,i}(t)$ is the population-level rate of acquiring serotype $i$ at time $t$ for an individual aged $a$ - which is all the positive terms in the ODE for $I_{a,i}$ - and dividing through by the population size of a given age ensures that we have a per capita rate.

Given that in 2001/2 the system could be considered to be in equilibrium, we applied an approximation using the equilibrium values,

$$Pr_{a,i}^{\text{inf}} \approx 1 - \exp\left(-(t_2 - t_1)\frac{T_{a,i}^*}{N_a}\right).$$

It should be noted that we do not have to include the probability of not having been infected up to sample time $t_1$, as this is included in the formulation of $T_{a,i}$.

We also know that the individual recovers between times $t_3$ and $t_4$. For a single infectious class, the recovery rate is $\gamma_{a,i}$. Hence, the probability of being infectious for time $\tau$ is $\gamma \exp(-\gamma\tau)$, giving

$$Pr_{a,i}^{\text{rec}} = \int_{t_1}^{t_2} \int_{t_3}^{t_4} \gamma_{a,i} \exp(-\gamma_{a,i}(s - t)) ds dt.$$

Equivalently,

$$Pr_{a,i}^{\text{rec}} = \frac{\exp(-\gamma_{a,i}(t_3 + t_4))[\exp(\gamma_{a,i}t_1) - \exp(\gamma_{a,i}t_2)][\exp(\gamma_{a,i}t_3) - \exp(\gamma_{a,i}t_4)]}{\gamma_{a,i}}.$$

Hence, the total probability for that positive individual was given by

$$Pr^{\text{pos}}_{a,i} = Pr^{\text{inf}}_{a,i} \times Pr^{\text{rec}}_{a,i}.$$

Note that if the first sample for the individual was positive we set $t_1 = t_2 - 30$, and if the last sample was positive we set $t_4 = t_3 + 365$.

A different calculation is needed if an individual fails to be recorded with serotype $i$ within the sample. This could be for two reasons: either the individual was never infected

$$Pr^{\text{never}}_{a,i} = \exp\left(-\int_{t_0}^{t_{end}} \frac{T_{a,i}(t)}{N_a} dt\right) \approx \exp\left(-(t_{end} - t_0)\frac{T^*_{a,i}}{N_a}\right)$$

or the individual was infected but recovered rapidly before being sampled:

$$Pr^{\text{rapid}}_{a,i} \approx \left\{1 - \exp\left(-(t_{end} - t_0)\frac{T^*_{a,i}}{N_a}\right)\right\} \int_0^{1 \text{ month}} \frac{1}{1 \text{ month}} \left\{1 - \exp(-\gamma_{a,i} t)\right\} dt$$

which we 'simplified' to

$$Pr^{\text{rapid}}_{a,i} \approx \left\{1 - \exp\left(-(t_{end} - t_0)\frac{T^*_{a,i}}{N_a}\right)\right\} \times \left\{1 - \frac{1 - \exp(-\gamma_{a,i} M)}{M\gamma_{a,i}}\right\}$$

where $M$ is one month.

Overall, the combined probability of these two events for an individual failing to be recorded with serotype $i$ within the sample period was

$$Pr^{\text{neg}}_{a,i} = Pr^{\text{never}}_{a,i} + Pr^{\text{rapid}}_{a,i}.$$

The log-likelihood of the data ($D$) given the parameters ($\theta$) was given by summing over all sampled individuals,

$$\log(L^{Carriage}_{2001}(D|\theta)) = \sum_{\text{positive for } i} \log(Pr^{\text{pos}}_{a,i}) + \sum_{\text{negative for } i} \log(Pr^{\text{neg}}_{a,i}).$$

#### S4.2 Sampling periods, beyond 2002

The second set of carriage data we used were cross-sectional samples taken in the years of 2008, 2012, 2015 and 2018 [4]. The sample dates for these data span a time period following the introduction of PCV7 (the 2008 sampling) to after the introduction of PCV13 (the 2012, 2015 and 2018 samplings).

During this time window there was no repeated sampling of the same individual. A possible issue to bear in mind is that for these sample periods we only get a snapshot for each individual and have no record of what other serotypes they may have caught during that time period (if any). In that aspect, the data gives a lower bound on number of carriage cases of a given serotype-age combination.

**Serotype-age group specific likelihood**
For the relevant time period for that particular round of sampling $y$, for each serotype $i$ and age group $a$, we computed a likelihood for the probability of getting the observed count of positive results $k_{i,a,y}$ given the total number of individuals sampled $n_{i,a,y}$ in that serotype-age group. i.e. a binomial distribution $X_{i,a,y} \sim B(n_{i,a,y}, p_{i,a,y})$.

In full,

$$Pr(X_{i,a,y} = k_{i,a,y}) = \binom{n_{i,a,y}}{k_{i,a,y}} p_{i,a,y}^{k_{i,a,y}} (1 - p_{i,a,y})^{n_{i,a,y} - k_{i,a,y}}$$

The probability of a 'successful' event $p_{i,a,y}$ was given by our ODE model estimate for average carriage prevalence for that serotype-age group during the time period $t$ under consideration (that is $p_{i,a,y} = Y \int_{t\in y} I_{a,i}(y)dt$, where $Y = \left[N_a \int_{t\in y} dt\right]^{-1}$). We approximated this value as $p_{i,a,y} = I_{a,i}(t)/N_a$ where $t$ is the mid-point of year $y$.

Thinking at an individual level, this is equivalent to the probability of a single, specific individual having carriage of the specified serotype at a randomly chosen time point during the time period under consideration.

**Likelihood for a single time period**
Now considering all serotype-age groups, the overall log-likelihood for a single sampling period $t$ was a product of probabilities from a set of binomial distributions (assuming independence of the binomial distributions from one another):

$$\log(L_t^{Carriage}(D|\theta)) = \sum_{i,a} \log(Pr(X_{i,a,t} = k_{i,a,t}))$$

where $t = \{2008, 2012, 2015, 2018\}$ .

**Likelihood across all time periods**
When considering all time periods, we could apply a weighting $w_t$ to each sample period:

$$\log(L^{Carriage}(D|\theta)) = \sum_{t} w_t \log(L_t^{Carriage}(D|\theta))$$

However, without any clear biases in sampling, we assumed $w_t = 1$.

Fig. S4 compares our model projections (lines) with the available carriage data (dots) for the five different sample times (2001/2, 2008, 2012, 2015 and 2018) [3, 4]. It is clear that despite the substantial efforts involved in collecting these data, the amount of carriage detected, especially in the older age groups, is limited.

### S5 Model fitting to IPD data

The available data from UKHSA was the number of reported IPD case count for the population for age-group $a$ (stratified into six groups: $< 2$, $2-4$, $5-14$, $15-44$, $45-64$ and 65+ years old), serotype $i$ and calendar year $y$ (from 2000 to 2023, inclusive): $C_{a,i,y}$.

We assumed that IPD cases were related to the modelled prevalence of pneumococcal carriage in the population ($I_{a,i}(t)$), with serotype and age dependent factors ($f_i$ and $F_a$) that included the need for hospital treatment and reporting. We also assumed that IPD cases were Poisson distributed:

$$C_{a,i,y}^{model} \sim \text{Poisson}\left(f_i F_a Y \sum_{X} \varrho^X \int_{t\in y} I_{a,i}^X(t)dt\right) = \text{Poisson}\left(M_{a,i,y}\right)$$

where $Y = \left[\int_{t\in y} dt\right]^{-1}$, to generate the correct daily average for modelled prevalence of pneumococcal carriage. We subdivide $I$ by the status of the individual when infected ($X = \{S, \text{PCV7}, \text{PCV13}, \text{PCV20}, \text{PPV23}\}$); and $\varrho^X$ (where $X$ is a vaccinated state) is the reduction in the conditional risk of disease (i.e. the risk of disease conditional on pneumococcal carriage). As such, the combined impact of reduced susceptibility to infection and reduced conditional risk of disease define the efficacy against disease for any given vaccine:

$$\text{Vaccine efficacy against disease}^X = VE_D^X = 1 - \rho^X \varrho^X$$

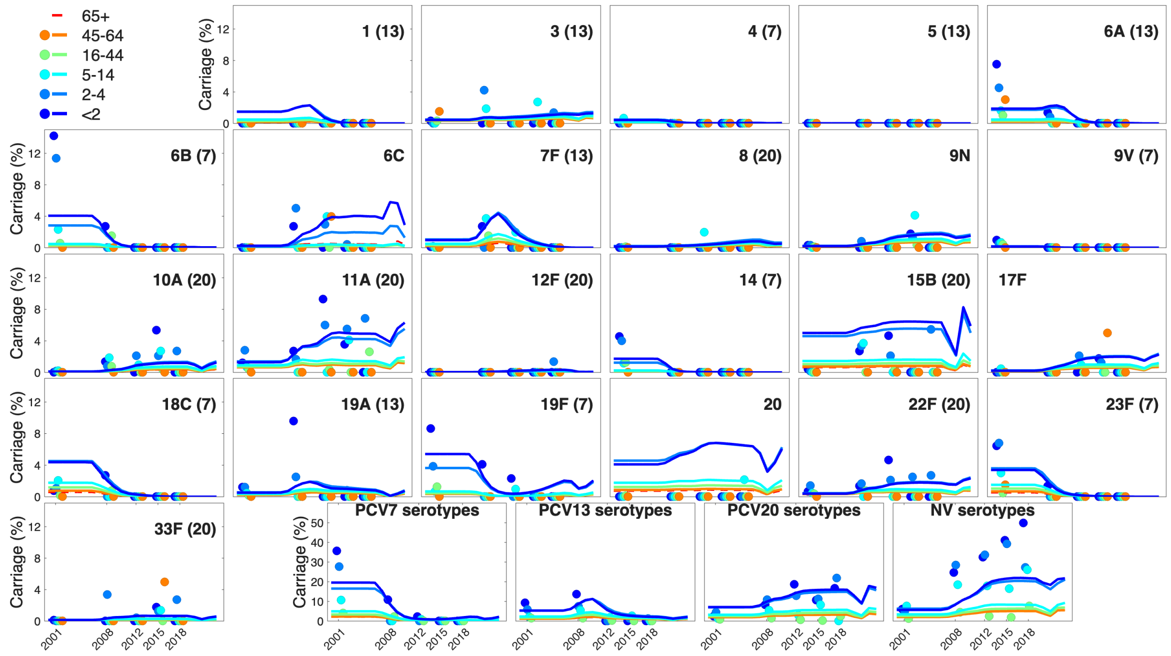

**Fig. S4. Match between best fit model results (lines) and data (dots) for age and serotype specific Carriage.** Colours denote the six different age groups. Serotype 2, which remains low throughout is not shown. The last four plots show the sum of serotypes in PCV7, PCV13 (but not PCV7), PCV20 (but not PCV13) and all other (non-vaccine) serotypes.

.

We worked with the resulting log-likelihood expression:

$$\log(L^{IPD}(C|\theta)) = \sum_{a,i,y} \left[C_{a,i,y} \log\left(M_{a,i,y}\right) - M_{a,i,y} - \log(C_{a,i,y}!)\right]$$

To reduce computational run time, we applied an approximation by defining a time $\tau_y$ mid-way through year $y$ where

$$Y \int_{t \in y} I_{a,i}(t)dt \approx I_{a,i}(\tau_y)$$

Overall, we defined the likelihood of observing the carriage and IPD data collectively given the model output as

$$\log(L^{Total}(C, D|\theta)) = \log(L^{IPD}(C|\theta)) + \sum_t \log(L^{Carriage}(D|\theta)).$$

Fig. S5 shows the results of this fitting process for IPD caused by 9 exemplar serotypes; showing examples of good agreement (Others, 6B, 9V, 11A and 33F), qualitative agreement (19F and 20) and poor agreement (1 and 18C). Note that, unlike the figures in the main paper, these comparisons are plotted using a linear scale which emphasises errors at larger values.

### S6 Model fitting to CAP data

We inferred an age distribution of pCAP from the data presented by Bewick *et al.* [6]. This study considered a sample of 920 patients with CAP in the Nottingham region over the years of 2008-2010,

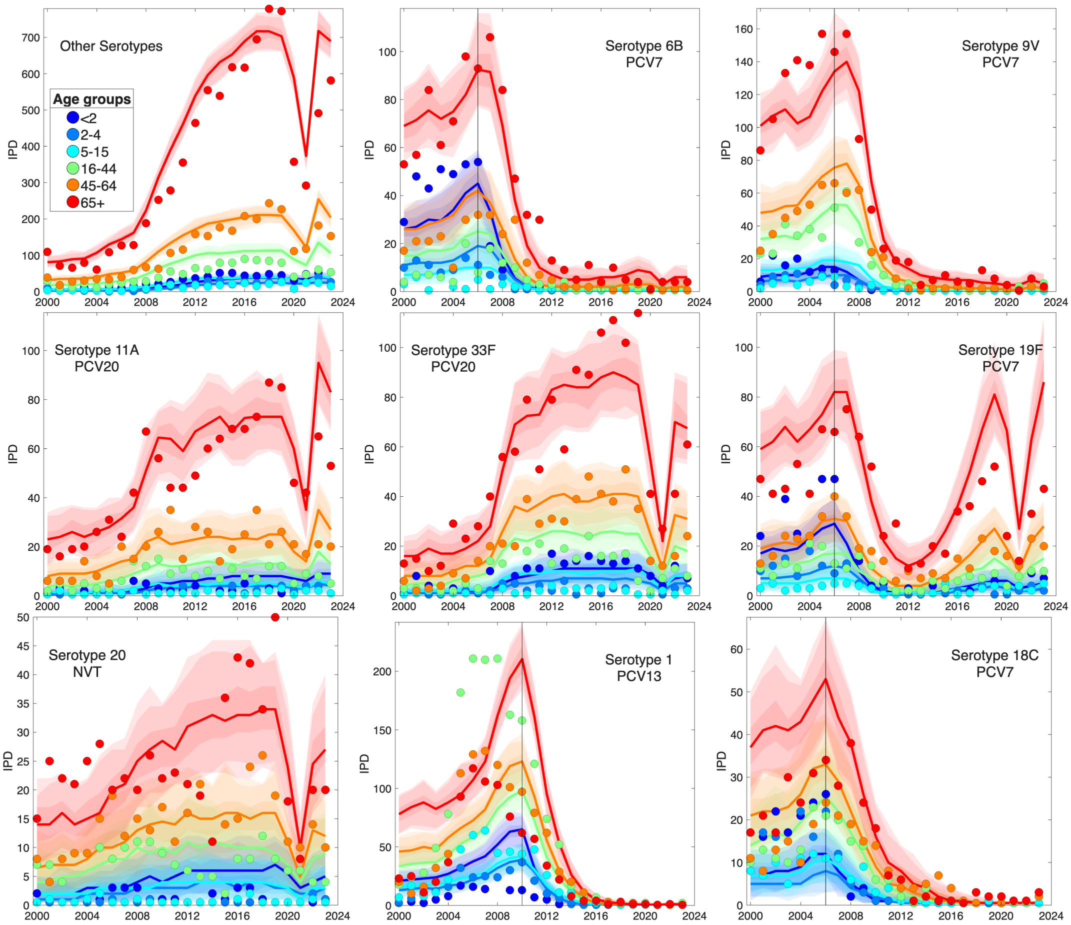

**Fig. S5. Match between model results (lines for the maximum likelihood estimates, and shaded ribbon for the prediction interval) and data (dots) for age and serotype specific IPD.** Colours denote the six different age groups: < 2 years (Dark Blue), 2-4 year olds (Blue), 5-14 year olds (Cyan), 15-44 year olds (Light Green), 45-64 year olds (Orange) and 65+ year olds (Red). Each serotype is labelled together with which vaccine it is included in. We show nine serotypes to highlight the range of behaviour. For the first 4 serotypes (Others, 6B, 9V, 11A and 33F) there is a good agreement between the model and the data. For the next 2 serotypes (19F and 20) the agreement is patchy, fitting the general qualitative trends but not the details. Finally the final two serotypes (1 and 18C) give examples of where the agreement is poor. For both of these serotypes we fail to capture the early age-structure: serotype 1, which is part of the PCV13 vaccine, has the peculiarity that it was most common in 15-44 year olds (green dots), hence while the model captures the general trend in incidence it cannot capture the age-dependence; IPD cases due to serotype 18C were relative common across all age-groups, again leading to the model over estimating the number of older cases.

among which 366 patients were identified with pCAP.

We inferred the serotype dependence of pCAP from the data studied by Lansbury *et al.* [5]. In this paper, they followed adults (those aged 16 years old and above) hospitalised with CAP between September 2013 and May 2014 in two teaching hospitals in Nottingham, except a pause during the COVID-19 pandemic between March and August 2020. A serotype-specific 24-valent urinary antigen assay was used to identify pneumococcal serotypes.

Of the 7078 patients diagnosed with CAP, 5186 gave their consent to participate in the study. The number of patients diagnosed with pneumococcal CAP was 2193 (42.3%). The trends of pCAP were given by year, by serotype, without age stratification. The maximum number of hospitalised pCAP cases was in 2018-2019, reaching 321 hospitalisations, while the lowest was during the pandemic year 2020-2021 with 125 admissions, followed by 2015-2016 with 165 admissions. Throughout the study period, serotype 3 was the most prevalent among hospitalised patients with 29% of the cases, followed by serotype 8 (21%), unassigned (8%), 15A (5%) and 9N (4%). However, these trends varied widely throughout the years. For example, serotype 3 steadily increased from 13% of hospitalisations in 2013-2014 to 49% in 2022-2023. Serotype 8 fluctuated from 17% in 2013-2014 to a maximum of 37% in 2020-2021 back to 20% in 2022-2023. These trends were made available in the supplementary material of Lansbury *et al.* [5]. We matched these adult trends to children aged 15 or less based on a 14.4 in $10,000$ incidence rate with CAP [7], out of which 46% are due to pneumococcal infection [8].

We performed this fitting in a manner similar to that of IPD. We assumed that pCAP was a Poisson sample of infected individuals with the rate determined by a combination of serotype and age dependent factors (denoted by $g_i$ and $G_a$, respectively):

$$\mathrm{pCAP}_{a,i,y} \sim \mathrm{Poisson}\left( g_i G_a Y \sum_X \varrho^X \int_{t \in y} I^X_{a,i}(t) dt \right)$$

We determined the parameters $g_i$ and $G_a$ by fitting to the Nottingham data. We then extrapolated to the national scale. We once more subdivide $I$ by the status of the individual when infected ($X = \{S, \mathrm{PCV7}, \mathrm{PCV13}, \mathrm{PCV20}, \mathrm{PPV23}\}$). Without additional information, we assumed the same reduction in the conditional risk of disease ($\varrho^X$) for both IPD and pCAP.

We note that given pCAP data was not taken from a representative national sample, we used only the data on IPD cases and carriage to fit the transmission model.

### S7 Health economic model parameterisation

#### S7.1 Cost per IPD hospitalisation

We obtained monthly hospitalisation data from the Hospital Episode Statistics (HES) database through the Department of Health. Data were available from 2003/2004 to 2015/2016. The cases were identified by the presence of the relevant diagnostic discharge codes listed in the patient's discharge record. The discrete health states or medical conditions associated with IPD were septicaemia (International Classification of Diseases, 10th revision [ICD-10], code (A403) and meningitis (G001); and for pCAP, we considered pneumonia due to Streptococcus pneumoniae (J13). Month and quinary age bands aggregated hospitalisation cases. Each observation comprised a Finished Consultant Episode (FCE), which measures the time the patient spends under the care of a particular consultant. We had episodes pertaining to each individual patient, which allowed us to construct a provider spell for each patient, measuring the time from admission to discharge. All successive episodes in each epidemiological year were linked.

We derived Healthcare Resource Group (HRG4) codes associated with diagnoses and treatment procedures for each FCE using the NHS Local Payment Grouper software. The grouper produces a single HRG4 code for every admission. It uses an algorithm that clusters diagnostic codes, treatments, procedures and length of stay (LoS) with similar resource implications. We mapped HRG4 codes to reimbursement costs by referring to the 2015/2016 NHS national tariff [22]. We used the same HRG grouping algorithm for data in all years. We adjusted the FCEs with excess bed days, not covered by the standard tariff (long stay trim point), using a per diem for each code.

We extracted data on surgical procedures and treatments that were identified through literature searches and expert opinion. To account for the differing degree of certainty in attributing hospital costs directly to meningitis and septicaemia, we categorised FCEs into the following six costing groups.

The primary diagnosis code was a condition associated with meningitis (G001) or septicaemia infection (A403) and:

- Group I: the grouping method used for the episode was driven by meningitis or septicaemia as the primary diagnosis.
- Group II: the grouping method used was driven by procedure/s compatible with the management of meningitis or septicaemia.
- Group III: the grouping method used was driven by procedure/s not directly attributable to meningitis or septicaemia.

The condition associated with meningitis or septicaemia infection is not a primary diagnosis and:

- Group IV: the grouping method used for the episode was driven by the primary diagnosis.
- Group V: the grouping method used was driven by procedure/s compatible with the management of meningitis or septicaemia as a secondary diagnosis.
- Group VI: the grouping method used was driven by procedure/s not directly attributable to meningitis or septicaemia.

Groups I, II and V give rise to costs that were directly a result of meningitis or septicaemia, whereas groups III, IV and VI give costs of hospital admissions due to other infections rather than meningitis or septicaemia. We undertook the analysis of costs annually and according to the following age groups: children (less than 2 years, 2-4 years and 5-14 years), working age adults (15-44 years and 45-64 years) and older adults (65-74 years, and 75+ years). We adjusted the costs to 2022/2023 prices using the Consumer Price Index (CPI) for Health provided by the Office for National Statistics [23].

##### S7.2 Cost per pCAP hospitalisation

The steps for deriving the costs of pCAP hospitalisations followed the same procedures as those for IPD. However, HRG4 codes were mapped to reimbursement costs using the latest 2022/2023 NHS national tariff [22]. Additionally, only cases with a primary diagnosis code of J13 were included in the analysis.

##### S7.3 QALY losses

**QALY loss per hospitalisation:** We sourced health state utility values from the literature. Due to limited evidence on the health-related quality of life (HRQoL) for individuals hospitalised with septicaemia, we collectively applied to IPD the utility value for meningitis hospitalisation [24]. Utility values for pCAP by age group were available only for adults aged 19 and above; therefore, we assumed that individuals under 19 had the same utility value as those aged 19–35 years [25].

To calculate the QALY loss per hospitalisation we performed the following three step process. We first estimated the average LoS for each age group for both IPD and pCAP. Second, we divided the inverse of the utility value for IPD or pCAP hospitalisation by 365, to obtain a QALY loss per day due to IPD or pCAP. Finally, we multiplied by the LoS for each respective age group, separately for IPD and pCAP, to arrive at our estimate for QALY loss per hospitalisation for IPD and pCAP.

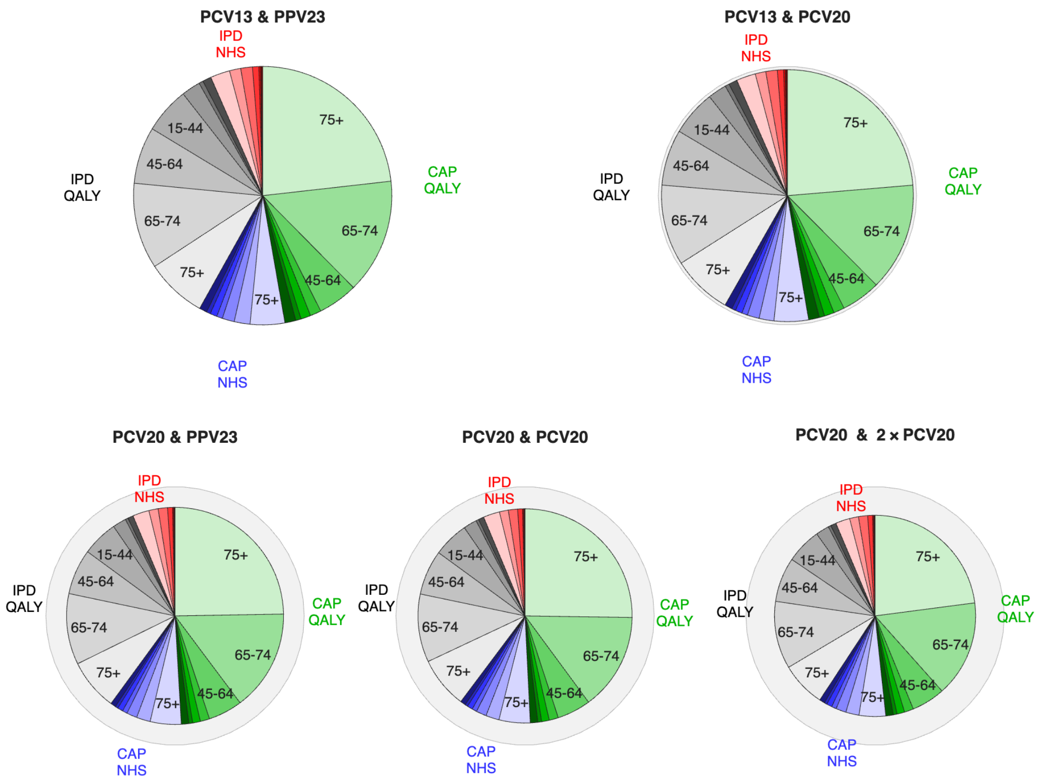

**Fig. S6. Cost components for the 50-year time horizon for differing vaccine valency combinations in pneumococcal vaccination programmes from 2026.** Displayed are age group stratifications of costs associated with IPD and CAP infection episodes (NHS healthcare costs and QALY losses at £20,000 per QALY) for four different scenarios for the vaccine used in the paediatric and adult pneumococcal vaccination programmes from 2026. The total areas of the pie charts are proportional to total cost. Listing the vaccine used in the paediatric programme first and adult programme second, the four scenarios shown are: (top left) PCV13 and PPV23; (top right) PCV13 and PCV20; (bottom left) PCV20 and PPV23; (bottom right) PCV20 and PCV20.

**QALY losses due to mortality:** We took year of age and sex stratified estimates of quality adjusted life expectancy for the population of England from McNamara *et al.* [26], with values discounted at a 3.5% annual discount rate. In brief, the production of the estimates reported by McNamara *et al.* [26] involved combining age- and sex-specific EQ-5D-5L utility scores (for 14,412 participants from the Health Survey for England in 2017 and 2018 [27, 28]) with national life tables of the English population (pooled for 2017-2019) [29]. To obtain our estimated QALY loss associated with mortality for each age group, we weighted the age and sex specific estimates for quality adjusted life expectancy from McNamara *et al.* [26] by the population structure for England recorded in the 2021 census [1].

**QALY losses due to sequelae:** We sourced QALY losses for individuals living with deafness, mild hearing loss, epilepsy, mild mental retardation, severe retardation and tetraplegia, and leg paresis from the literature. We report risks on a per-year basis [30] (Table S2).

#### S7.4 Health Economics and Long-term projections

By combining the long-term model projections together with the health economic calculations for direct healthcare costs and QALY losses, we evaluate four different vaccination options for 2026 (**??**). These results can be compared to Fig. 4, but show the costs across a 50-year time horizon with

**Table S2. Risk of sequelae from meningitis and associated proportional reduction in QALYs per year of life**. All risks are assumed to be age-independent and are reported on a per-year basis [43]. We report all values to 2 s.f.

| Sequelae | Risk | QALY reduction |
|---|---|---|
| Deafness | 10% | 53% |
| Mild hearing loss | 34% | 26% |
| Epilepsy | 6.5% | 22% |
| Mild mental retardation (MR) | 4.2% | 56% |
| Severe retardation and tetraplegia | 3.1% | 98% |
| Leg paresis | 8.7% | 36% |

discounting (at 3.5%). We note that while the switch from PCV13 in infants and PPV23 in older adults to PCV20 in both is associated with a decline in the overall costs (reducing from an average of £333M per year to £263M), there is relatively little change in how this cost is distributed between age groups or causes.

An alternative way to visualise these results is by presenting both projections of IPD and projections of total costs (including IPD and pCAP); see Fig. S7, which can be compared to Fig. 1. In all cases, disease in the older age-group (65+) dominates the patterns.

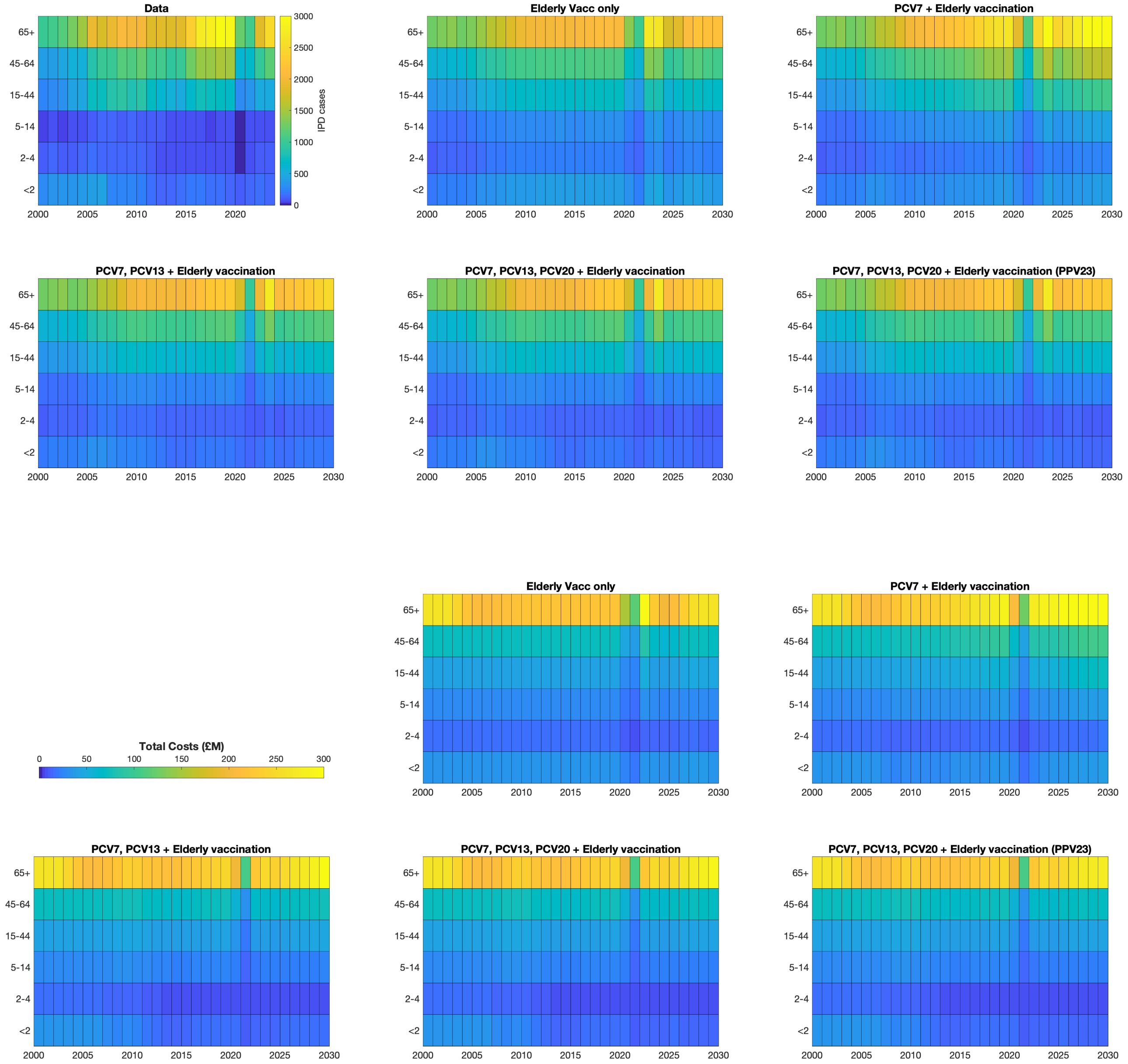

**Fig. S7. Impact of different vaccination programmes on IPD and pneumococcal costs.** Five different vaccination scenarios are considered, in the first four there is a switch from PPV23 to PCV20 vaccination in the elderly in 2026, in the last one PPV23 vaccination in continued. The paediatric vaccination programmes are: none; PCV7 only from 2006; PCV7 in 2006 and PCV13 from 2010; and PCV7 in 2006, PCV13 in 2010 and PCV20 from 2026 onwards. The upper panel shows the number of IPD cases in each age group over time, from the data (top left, note the shorter time-axis) and from model simulations with different vaccination assumptions. The lower panel shows the estimated annual cost. We amalgamated the 65-74 and 75+ age groups into a single class to aid comparisons, though we ran the dynamics and calculated the costs treating them as two distinct two age groups.

#### S8 Statistical fits to IPD data

To further support our analysis, here we perform simple statistical fits to the IPD data for each serotype (noting that the proportion of IPD cases that are serotyped has changed over time from around 50% in the early 2000s to around 95% at the time of writing).

We assume that the dynamics of each serotype in each age group can be fit by a series of sigmoidal growth (or decay) curves. Specifically, we assumed that the number of cases of IPD in a given year, $y$, was Poission distributed, accounting for the mean dynamics ($M_{a,s}$) and the proportion of cases that are serotyped ($P_y$):

$$\mathit{IPD}(y, a, s) \sim \text{Poisson}\left(P_y M_{y,a,s}\right)$$

with $M$ being a piecewise function, with three piecewise elements spanning 2000-2005, 2006-2009 and 2010-2019, chosen such that they span pre-vaccination, vaccination with PCV7 and vaccination with PCV13.

$$M_{y,a,s} = \begin{cases} B_0 & \text{if } y \leq 2005 \\ A_1 + (B_1 - A_1)\exp(r_1(y - Y_1))/(1 + \exp(r_1(y - Y_1))) & \text{if } 2006 \leq y \leq 2009 \\ A_2 + (B_2 - A_2)\exp(r_2(y - Y_2))/(1 + \exp(r_2(y - Y_2))) & \text{if } 2010 \leq y \leq 2019 \end{cases}$$

where we defined the values of $A_1$ and $A_2$ so that individual elements agreed in 2006 and 2010 (Fig. S8). We note that no attempt is made to fit to COVID-19 era data. We estimated all parameters with a simple Metropolis-Hastings MCMC scheme, and weak uninformative priors are used throughout. (We use one million iterations of the MCMC scheme, after a burn-in period of a million iterations; our priors are $r_i \in U(0, 1)$, $Y_i \in U(2005, 2019)$, $B_i \in \text{Exp}(Z_{a,s}^{-1})$ where $Z_{a,s} = 2\max_y\left[\mathit{IPD}(y, a, s)\right]$.) The red curve (which is $P_y M_{y,a,s}$) is in good agreement with the data.

By comparing the black, green and blue lines in Fig. S8 (which correspond to no paediatric vaccination, PCV7 and PCV13), and incorporating the health economic costs of IPD, we can obtain an estimate of the Willingness to Pay (WTP) Thresholds for PCV7 and PCV13. We computed the WTP thresholds over a 50-year time horizon and only included (health and healthcare) costs associated with IPD (Fig. S9). These simple estimations compare well with the more complex ODE projections. In summary, it suggests that vaccination using PCV7 was not cost effective due to serotype replacement, but a switch to PCV13 was likely to be cost effective. The simple model underestimates the willingness to pay threshold compared to the fully ODE model for PCV13, but both modelling approaches estimate that a switch from PCV7 to PCV13 will be cost effective if the vaccine price is sufficiently low.

Unfortunately, this approach cannot be applied to the paediatric introduction of PCV20, given the absence of data on its population-level impacts. However, this approach does offer a fast method of readily gauging the long-term impact in the first few years following introduction of a new scheme.

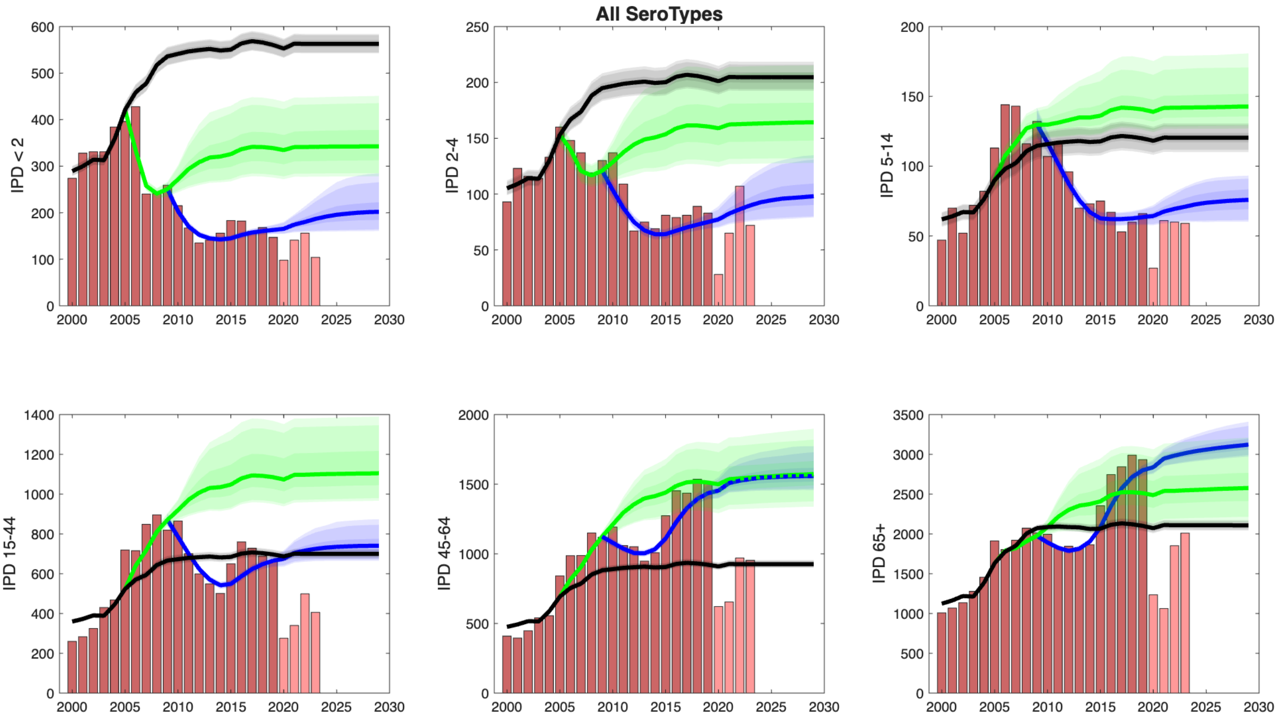

**Fig. S8. Sum of the fits to individual serotypes by age**. Bars show the reported, serotyped IPD cases, with darker bars showing the years used in the fitting. Following the colour scheme of Fig. 6b, black lines show the extrapolation of pre-vaccination levels (account for changes in serotype detection); green lines correspond to PCV7 (2006-2010) and its extrapolation into the future; blue lines correspond to PCV13 (from 2010 onwards). The shaded regions show the prediction interval due to parameter uncertainty.

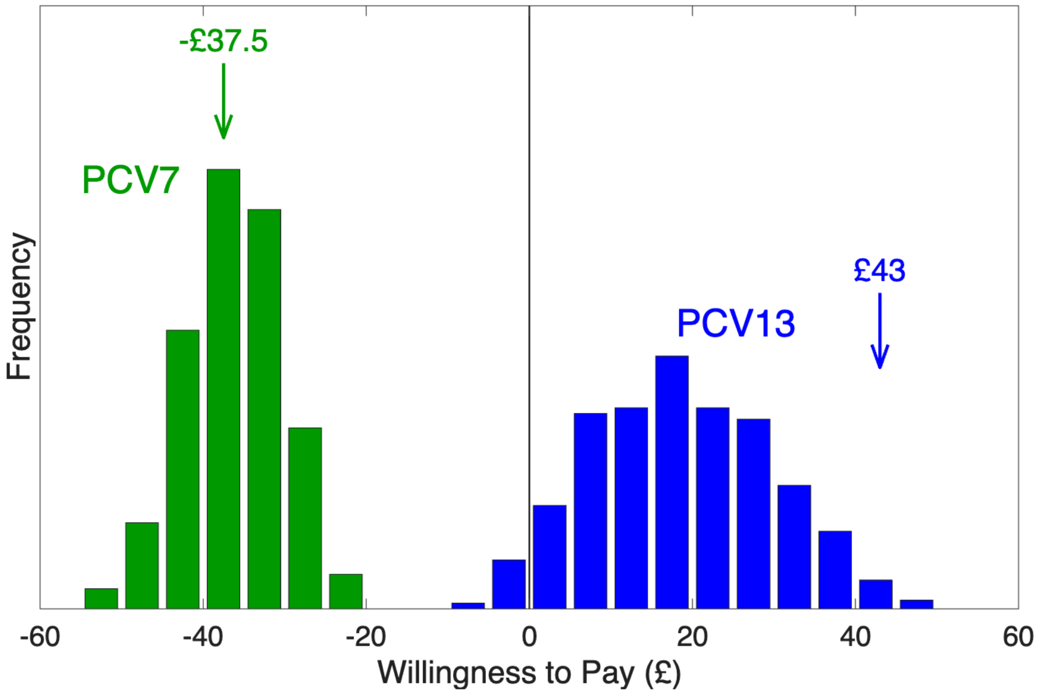

**Fig. S9. Willingness to Pay Thresholds from the simple statistical model**. Green represents the WTP for the PCV7 vaccine in 2006, assuming three doses and a £10.50 administration charge. Blue represents the WTP for replacing PCV7 with PCV13. All results are over a 50-year time horizon with 3.5% discounting, and only include costs due to IPD. Arrows represent the central WTP estimates from the full ODE model (only including IPD associated costs), distributions reflect the parameter uncertainty from the statistical fitting.
